# When Fairness Metrics Mislead: Demographic Parity Reduces HIV Screening for High-Burden Populations

**DOI:** 10.64898/2026.01.27.26344936

**Authors:** Hayden Farquhar

## Abstract

**Background:** In clinical contexts where disease burden differs across demographic groups, enforcing demographic parity — equal prediction rates regardless of group — may reduce screening for the populations that need it most. We demonstrate this using HIV testing prediction as a case study.

**Methods:** Using the Behavioral Risk Factor Surveillance System (BRFSS) 2024 dataset (N=386,775), we trained four classifiers to predict HIV testing uptake and evaluated disparities using demographic parity difference (DPD), equalized odds difference (EOD), and calibration across eight racial/ethnic groups. We applied threshold optimization and exponentiated gradient mitigation and quantified their impact on high-burden populations, including intersectional effects across race and sex.

**Results:** Baseline selection rates ranged from 12.1% (Asian) to 66.0% (Black), mirroring differential HIV burden (DPD 0.519-0.634). Race-blind models retained 70% of baseline disparity through correlated social determinants. Enforcing demographic parity reduced Black true positive rates from 78.2% to 30.0% (61.6% relative decrease), causing 1,610 additional missed individuals. Race-only optimization worsened sex-based disparity by 71%; multi-objective optimization reduced intersectional DPD from 0.609 to 0.076 but at the same cost to high-burden groups. Exponentiated gradient AUC fell from 0.671 to 0.592 (11.8% relative decrease). Survey-weighted sensitivity analysis confirmed unweighted estimates underestimated disparities.

**Conclusions:** Demographic parity is an inappropriate fairness criterion in differential-burden clinical contexts because it reduces screening access for high-risk populations. Fairness audits in healthcare should use need-appropriate metrics (equalized odds, calibration) rather than defaulting to demographic parity, and metric selection should involve clinician and community stakeholder deliberation.

## Introduction

### Algorithmic Fairness in Healthcare: A Growing Imperative

Algorithmic fairness — whether predictive models perform equitably across demographic groups — is now a standard consideration in healthcare machine learning (ML) (1,2). High-profile examples have catalyzed attention: a widely deployed commercial algorithm for identifying high-risk patients was found to systematically disadvantage Black patients, requiring 2.5 times higher predicted risk to receive equivalent care recommendations (3). Fairness audits have since documented disparities in dermatology AI (4), sepsis prediction (5), and cardiovascular risk scoring (6). Frameworks for quantifying and mitigating algorithmic bias are now well established (7,8).

But one question remains underexamined: **which fairness metric should be applied, and when does the choice of metric itself cause harm?**

### The Metric Selection Problem

The algorithmic fairness literature offers multiple, mathematically incompatible definitions of fairness (9,10). Two dominant paradigms illustrate the tension:

**Demographic parity** requires equal positive prediction rates across groups regardless of outcome rates. This is appropriate when group membership is irrelevant to the outcome — for example, loan approval, where race should not influence creditworthiness assessment.

**Equalized odds** requires equal true positive and false positive rates across groups, conditioning on the actual outcome. This is appropriate when base rates legitimately differ across groups — the model should be equally accurate for all groups, but need not select at equal rates.

**Calibration** requires that predicted probabilities carry the same meaning across groups — among individuals predicted to have 70% probability, approximately 70% should truly be positive regardless of group membership.

These metrics cannot all be satisfied simultaneously when base rates differ across groups (9). In healthcare, base rates — disease prevalence, risk factor distributions, care utilization patterns — frequently differ across demographic groups due to genuine epidemiological variation, structural inequities, or both. The choice among fairness metrics is therefore not merely technical but normative, with direct consequences for patient care.

### HIV as a Critical Test Case

Human immunodeficiency virus (HIV) sharply illustrates this problem. HIV burden in the United States is profoundly unequal: Black/African American individuals constitute 13% of the population but account for 42% of new diagnoses; Hispanic/Latino individuals represent 19% of the population but 27% of new diagnoses (11). Black men who have sex with men (MSM) experience the highest incidence of any demographic group (11,12). These disparities reflect structural factors — poverty, insurance gaps, geographic barriers, stigma, and the legacy of systemic racism (12–14) — not biological differences.

ML prediction models are increasingly deployed across the HIV care continuum: identifying candidates for pre-exposure prophylaxis (PrEP) (15,16), predicting viral suppression (17,18), forecasting care retention (19,20), and optimizing prevention resource allocation (21,22). Our systematic search of PubMed (search strategy detailed in Supplementary Materials, Section 6) identified 9,201 articles related to HIV prediction and machine learning published between 2017 and 2026, with 259 specifically addressing ML-based HIV prediction models — yet we identified no comprehensive fairness audit using standard algorithmic fairness metrics.

If demographic parity is applied to HIV testing models — requiring equal testing recommendation rates across racial groups — it would force models to *reduce* screening recommendations for high-burden populations (Black, Hispanic) and *increase* them for lower-burden populations (White, Asian). This represents a potential inversion of public health priorities: achieving “statistical fairness” by reducing access for those with the greatest clinical need.

### Study Objectives

We conducted a fairness analysis of HIV testing prediction models to demonstrate the consequences of fairness metric selection in a differential-burden context. Our objectives were to:

1. Quantify differential prediction rates across racial/ethnic groups in standard ML classifiers trained on nationally representative data
2. Demonstrate that race-correlated features perpetuate differential prediction even in race-blind models
3. Show that demographic parity optimization reduces screening recommendations for high-burden populations, potentially causing clinical harm
4. Characterize the impact of fairness optimization across intersectional groups (race x sex) and demonstrate cross-dimensional trade-offs
5. Argue that equalized odds and calibration-based metrics are more appropriate fairness criteria in differential-burden healthcare contexts

## Methods

### Study Population and Data Sources

#### Primary Analysis Dataset: BRFSS 2024

We used the Behavioral Risk Factor Surveillance System (BRFSS) 2024 dataset as our primary data source. BRFSS is a nationally representative telephone survey of U.S. adults conducted annually by the CDC, collecting data on health-related risk behaviors, chronic health conditions, and use of preventive services. The 2024 survey included 457,670 respondents across all 50 states and U.S. territories.

From the full survey, we identified 393,676 respondents (86.0%) with valid HIV testing responses. After excluding records with missing demographic information (race/ethnicity, sex, age, region), our final analysis dataset comprised 386,775 adults. After applying additional feature completeness requirements (non-missing values for all predictor variables), the modeling dataset comprised 323,924 adults, which was split into training (N=259,139; 80%) and test (N=64,785; 20%) sets using stratified random sampling to preserve outcome and demographic distributions. The full analysis sample of 386,775 was retained for descriptive statistics and survey-weighted sensitivity analyses.

The primary outcome was HIV testing behavior, defined as ever having been tested for HIV (HIVTST7 variable). Overall, 36.4% of respondents reported prior HIV testing. This outcome captures *who has been tested* — a measure of historical healthcare utilization — rather than *who should be tested* based on clinical risk. This distinction is central to our analysis: a model trained on testing uptake learns existing care-seeking and public health targeting patterns, including any biases embedded in those patterns. We examine what happens when fairness metrics are applied to such a model without accounting for this distinction.

Predictor variables included: age (continuous), sex (male/female), race/ethnicity (8 categories: White, Black, Hispanic, Asian, American Indian/Alaska Native [AIAN], Native Hawaiian/Pacific Islander [NHPI], Multiracial, Other), low income status (household income <$25,000), depression diagnosis (ever told by provider), cost barrier to care (could not see doctor due to cost in past 12 months), poor self-rated health status, and geographic region (South vs. Non-South).

The demographic composition of the analysis dataset was: White (73.4%), Hispanic (10.8%), Black (7.8%), Asian (2.8%), Multiracial (2.3%), AIAN (1.4%), Other (0.8%), NHPI (0.5%). HIV testing rates varied substantially by race/ethnicity: Black (56.3%), AIAN (48.9%), Multiracial (48.8%), Hispanic (47.3%), Other (44.4%), NHPI (33.3%), White (32.5%), Asian (24.5%).

#### Supplementary Demographic Data: NHANES 2017-2018

We obtained demographic and health insurance data from the National Health and Nutrition Examination Survey (NHANES) 2017-2018 cycle (N=9,254 respondents). NHANES provided additional context on demographic distributions, health insurance coverage, depression screening (PHQ-9), and general health status patterns used to validate BRFSS variable coding.

#### External Validation Data

**Ryan White HIV/AIDS Program Data:** We obtained aggregate viral suppression rates by race/ethnicity from the 2024 Ryan White HIV/AIDS Program Annual Client-Level Data Report (N=372,220 patients served). This dataset represents the largest federally funded HIV care program in the United States and provides ground truth on clinical outcomes by demographic group, enabling us to assess whether differential prediction rates in our models correspond to genuine differential burden.

**CDC AtlasPlus:** We extracted state-level HIV surveillance data (N=36,190 records across 13 indicator files) to examine geographic variation in racial disparities. Data included viral suppression rates, new diagnoses, and care continuum metrics by race/ethnicity and state.

**AIDSVu:** We obtained county-level HIV prevalence and care continuum data (80 files) from Emory University to contextualize geographic patterns in disparities.

### Coefficient Audit of Published HIV Risk Scores

To examine explicit race-based risk stratification in existing tools, we reviewed the coefficients of published HIV risk scores:

**Denver HIV Risk Score.** The Denver HIV Risk Score (23) is a validated point-based instrument for identifying individuals at elevated HIV risk in emergency department settings. The score assigns points for: age 22-25 years (+4), age 26-32 (+6), age 33-46 (+4), male sex (+8), **Black race (+7)**, **Hispanic ethnicity (+3)**, men who have sex with men (+10), injection drug use (+9), and vaginal sex without condom (+5). The score explicitly includes race as a predictor, with Black patients receiving +7 points regardless of behavioral risk factors.

*Note: The Denver Score requires behavioral variables (MSM status, injection drug use, condom use) not available in BRFSS. We present the Denver Score as a coefficient audit demonstrating explicit race-based risk stratification, rather than a performance evaluation on our dataset*.

### Machine Learning Classifiers

We trained four standard ML classifiers to predict HIV testing status:

- **Logistic Regression (LR):** L2-regularized logistic regression with hyperparameter tuning via 5-fold cross-validation
- **Random Forest (RF):** Ensemble of 100 decision trees with max depth 10, minimum samples per leaf 5
- **Gradient Boosting (GB):** Gradient boosted trees with learning rate 0.1, max depth 5, 100 estimators
- **XGBoost (XGB):** Extreme gradient boosting with similar hyperparameters, optimized for performance

Feature sets included 8 variables: age, sex (male), race_black, race_hispanic, low_income, depression_dx, cost_barrier, and poor_health. For race-blind modeling experiments, we excluded race variables and trained on the remaining 6 features to test whether differential prediction persists through correlated features.

### Fairness Metrics

We evaluated model fairness using three established metrics from the algorithmic fairness literature (3,7), deliberately comparing their implications:

**Demographic Parity Difference (DPD):** Measures the difference in positive prediction rates (selection rates) between the most and least selected groups: DPD = max(P(Y_hat=1|A=a)) - min(P(Y_hat=1|A=a)) across all protected attribute values a. A DPD of 0 indicates equal selection rates. DPD does not condition on the outcome and assumes equal selection rates are desirable — an assumption we critically examine.

**Equalized Odds Difference (EOD):** Measures the maximum difference in true positive rates (TPR) and false positive rates (FPR) across groups: EOD = max(|TPR_a - TPR_b|, |FPR_a - FPR_b|) for all group pairs (a,b). EOD conditions on the actual outcome and captures whether model accuracy is equivalent across groups, independent of base rates.

**Calibration:** Whether predicted probabilities carry the same meaning across groups, assessed via calibration curves and Brier scores stratified by race/ethnicity.

We also computed:

**True Positive Rate Gap:** The difference in TPR between specific groups (e.g., TPR_Black - TPR_White), indicating whether positive cases are equally likely to be identified across groups.

**Cohen’s h Effect Size:** The standardized difference in proportions (selection rates) between groups, with |h|<0.2 considered small, 0.2-0.8 medium, and >0.8 large.

Following established conventions, we considered |DPD| > 0.1 and |EOD| > 0.1 as exceeding acceptable fairness thresholds, though we note that appropriate thresholds depend on clinical context and stakeholder values (9).

### Fairness Mitigation Methods

We implemented two categories of fairness interventions, deliberately optimizing for demographic parity to demonstrate its consequences:

**Threshold Optimization (Post-processing):** This method (24) finds group-specific classification thresholds that optimize a fairness criterion while preserving the underlying model’s predicted probabilities. We optimized for demographic parity, selecting thresholds that equalized positive prediction rates across racial groups.

**Exponentiated Gradient (In-processing):** This method (25) incorporates fairness constraints directly into model training via Lagrangian-style optimization. The algorithm iteratively adjusts sample weights to satisfy fairness constraints while minimizing classification loss. We used L2-regularized logistic regression as the base estimator for exponentiated gradient (as required by the reductions framework), and tested demographic parity and equalized odds constraints across a range of tolerance parameters (epsilon = 0.001 to 0.2) to characterize the accuracy-fairness Pareto frontier. Performance comparisons between exponentiated gradient and XGBoost baseline therefore reflect both the fairness constraint and the change in base model complexity.

### Statistical Analysis and Robustness

Model performance was assessed using area under the receiver operating characteristic curve (AUC), accuracy, precision, recall, F1 score, and Brier score. Fairness metrics were computed using the Fairlearn library. Statistical significance of selection rate differences was assessed using two-proportion z-tests with Bonferroni correction for multiple comparisons (alpha = 0.0167 for three pairwise comparisons).

Robustness was assessed through multiple approaches:

- Bootstrap resampling (500 iterations) to construct 95% confidence intervals for DPD estimates
- 5-fold cross-validation (N=64,785 per fold) to assess consistency across data splits
- Sensitivity analysis across 10 random seeds to assess stability
- Sensitivity analysis across different base classifiers (LR, RF, GB, XGBoost)
- Sensitivity analysis across constraint tolerance parameters (epsilon from 0.001 to 0.2)
- McNemar’s test to assess whether predictions differed significantly before versus after mitigation
- Chi-square tests for independence between predictions and race

Clinical utility was assessed using decision curve analysis (26), comparing net benefit of baseline versus mitigated models across threshold probability ranges.

### Survey Design Considerations

BRFSS uses a complex survey design with stratification, clustering, and oversampling of certain populations. Our primary analyses treat the sample as unweighted. We acknowledge this as a limitation: unweighted estimates may not reflect nationally representative prevalence or disparity estimates. However, our primary research question concerns the *relative behavior* of fairness metrics and mitigation methods, which is less sensitive to survey weighting than point prevalence estimation. We discuss the implications of this limitation and the need for weighted replication in the Limitations section.

All analyses were conducted in Python 3.12 using scikit-learn 1.8.0, Fairlearn 0.13.0, XGBoost 3.2.0, and custom analysis pipelines.

## Results

### Baseline Model Performance

All four ML classifiers achieved moderate discrimination in predicting HIV testing status (Table 1). AUC ranged from 0.671 (Logistic Regression) to 0.714 (Gradient Boosting).

**Table 1.** Baseline Model Performance (Test Set, N=64,785)

| Model | AUC | Accuracy | Precision | Recall | F1 | DPD | EOD |
| --- | --- | --- | --- | --- | --- | --- | --- |
| Logistic Regression | 0.671 | 0.649 | 0.571 | 0.293 | 0.387 | 0.634 | 0.669 |
| Random Forest | 0.714 | 0.677 | 0.622 | 0.377 | 0.469 | 0.579 | 0.627 |
| XGBoost | 0.713 | 0.676 | 0.613 | 0.392 | 0.478 | 0.539 | 0.586 |
| Gradient Boosting | 0.714 | 0.679 | 0.621 | 0.391 | 0.480 | 0.519 | 0.565 |

All models exhibited substantial differential prediction rates. DPD ranged from 0.519 to 0.634, and EOD ranged from 0.565 to 0.669 — both 5-6 times higher than the conventional 0.1 threshold. Notably, DPD and EOD were similar in magnitude, reflecting the close relationship between selection rate differences and accuracy differences when base rates vary.

### Differential Prediction Rates by Race/Ethnicity

Table 2 presents baseline performance stratified by race/ethnicity for the XGBoost classifier.

**Table 2.** XGBoost Baseline Performance by Race/Ethnicity.

| Race | N | Testing Rate | Selection Rate | TPR | FPR |
| --- | --- | --- | --- | --- | --- |
| White | 48,173 | 34.0% | 16.3% | 28.2% | 10.1% |
| Black | 4,901 | 57.8% | 66.0% | 79.0% | 48.2% |
| Hispanic | 6,733 | 48.9% | 52.7% | 63.7% | 42.3% |
| Asian | 1,736 | 27.2% | 12.1% | 20.3% | 9.0% |
| AIAN | 932 | 51.1% | 28.5% | 37.4% | 19.3% |
| NHPI | 278 | 37.4% | 18.3% | 23.1% | 15.5% |
| Multiracial | 1,569 | 48.7% | 27.8% | 39.7% | 16.5% |
| Other | 463 | 45.8% | 21.8% | 28.3% | 16.3% |

Selection rates ranged from 12.1% (Asian) to 66.0% (Black) — a 5.5-fold difference ( $z=81.52$ , $p<0.001$ for Black-White comparison). However, a critical observation emerges when comparing selection rates to actual testing rates: the rank ordering closely mirrors actual HIV testing prevalence. Groups with higher testing rates (Black: 57.8%, Hispanic: 48.9%) received higher selection rates, while groups with lower testing rates (Asian: 27.2%, White: 34.0%) received lower selection rates. This pattern is consistent with a model that has learned to predict the outcome it was trained on — testing utilization — rather than evidence of algorithmic malfunction.

**Figure 1.**
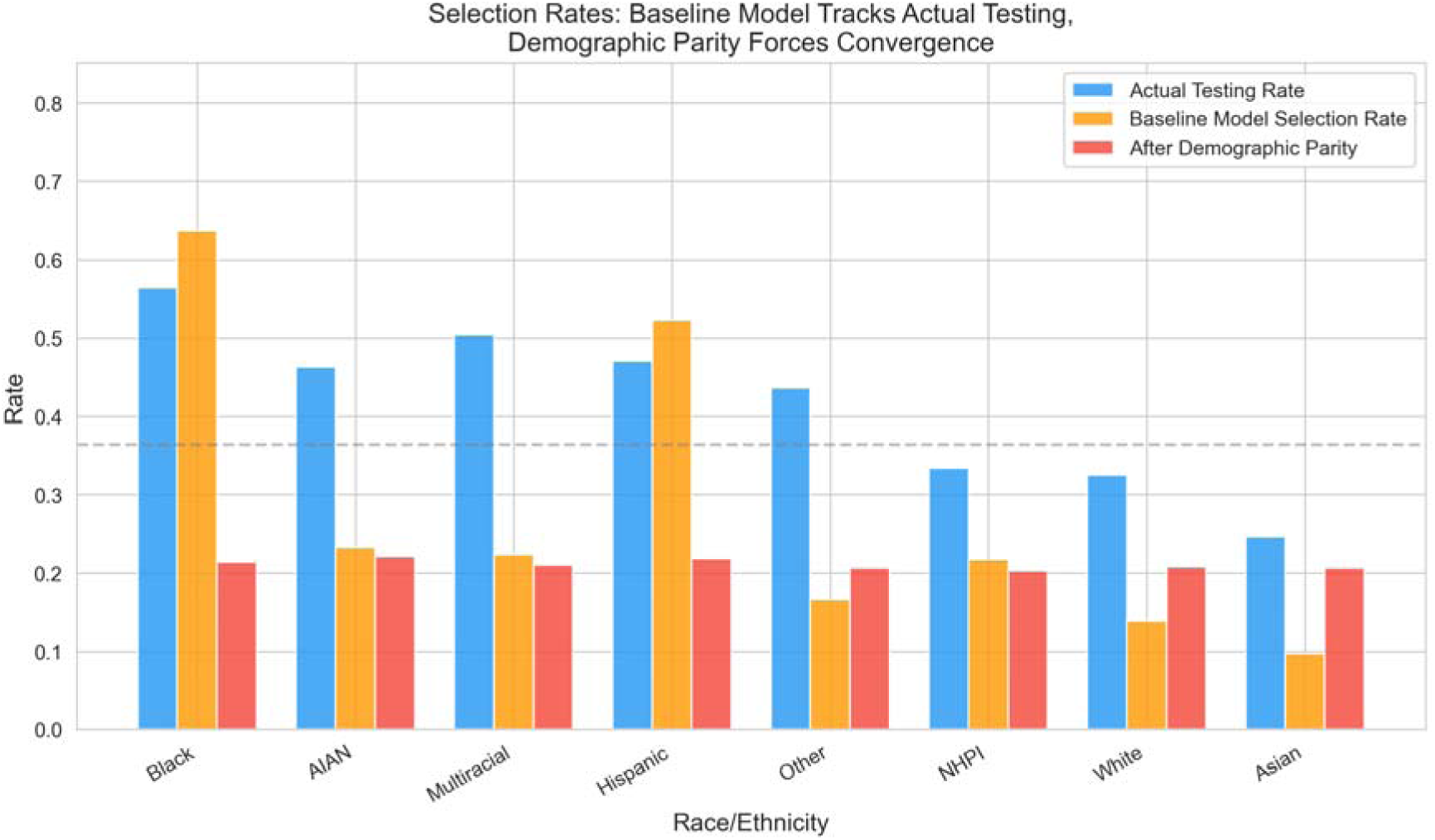
Comparison of actual HIV testing rates, baseline model selection rates, and demographic parity-mitigated selection rates by race/ethnicity. Baseline selection rates closely mirror actual testing prevalence, while DP mitigation compresses all groups toward a uniform rate, substantially reducing selection for high-burden groups (Black, Hispanic) while increasing it for lower-burden groups (White, Asian).

### Race-Blind Models: Persistent Differential Prediction Through Correlated Features

Race-blind models (excluding race variables) retained substantial differential prediction rates (Table 3).

**Table 3.** Race-Inclusive vs. Race-Blind Model Comparison.

| Model | DPD | EOD | AUC | Disparity Retained |
| --- | --- | --- | --- | --- |
| LR (with race) | 0.634 | 0.669 | 0.671 | — |
| LR (no race) | 0.154 | 0.186 | 0.656 | 70.1% |
| XGB (with race) | 0.539 | 0.586 | 0.713 | — |
| XGB (no race) | 0.169 | 0.166 | 0.702 | 68.7% |

Removing race from the feature set eliminated only 30% of differential prediction. Approximately 70% of baseline disparity persisted through race-correlated features: insurance status, depression diagnosis, cost barriers to care, and geographic region (South vs. Non-South). These variables are themselves shaped by structural inequities — the same socioeconomic factors that drive differential HIV burden also drive differential values on these predictors. The model captures real associations in the data: individuals with cost barriers, low income, and Southern residence are both more likely
to have been tested for HIV (through targeted public health campaigns) and more likely to be Black or Hispanic.

This finding has two distinct interpretations depending on the clinical context:

- **If the model is used to predict who needs testing:** Persistent differential prediction through social determinants may represent appropriate capture of risk-correlated factors
- **If the model is used to allocate scarce resources equitably:** Persistent differential prediction indicates that removing race alone is insufficient to achieve equitable allocation

### Calibration Analysis

Calibration varied across racial groups (Table 4).

**Table 4.**
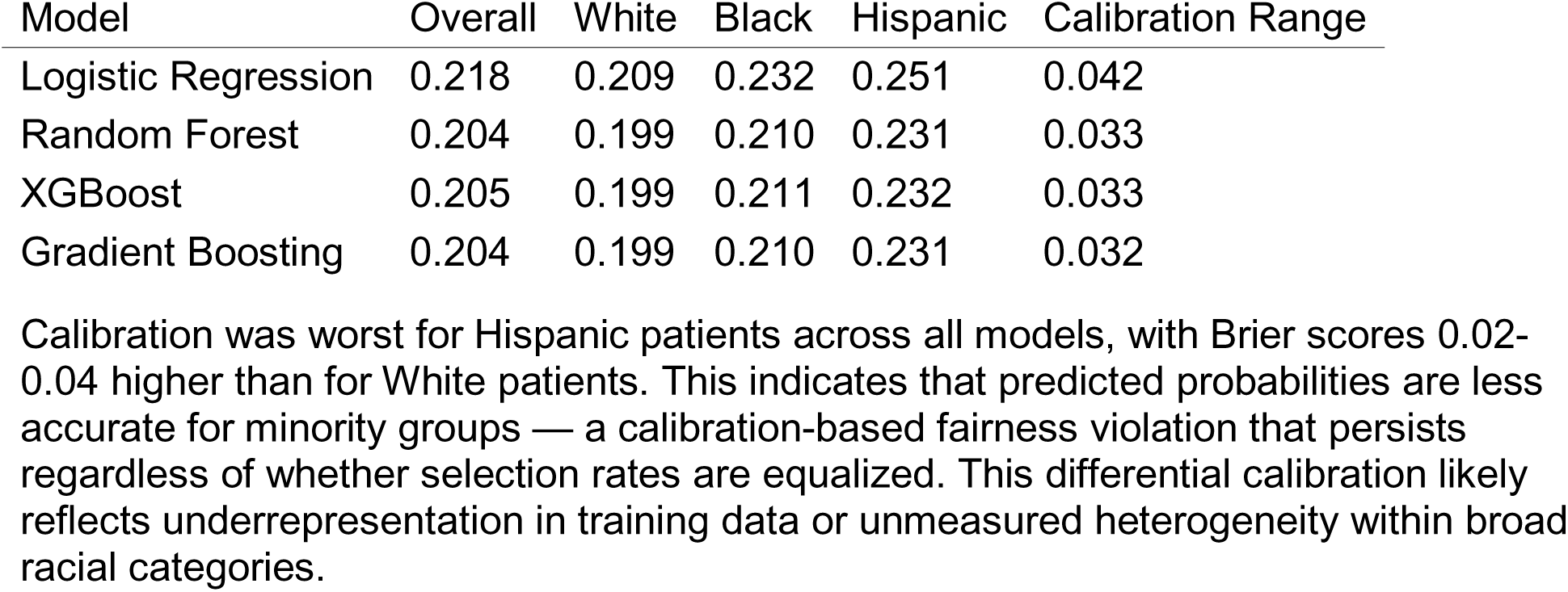
Brier Scores by Race and Model.

| Model | Overall | White | Black | Hispanic | Calibration Range |
| --- | --- | --- | --- | --- | --- |
| Logistic Regression | 0.218 | 0.209 | 0.232 | 0.251 | 0.042 |
| Random Forest | 0.204 | 0.199 | 0.210 | 0.231 | 0.033 |
| XGBoost | 0.205 | 0.199 | 0.211 | 0.232 | 0.033 |
| Gradient Boosting | 0.204 | 0.199 | 0.210 | 0.231 | 0.032 |

Calibration was worst for Hispanic patients across all models, with Brier scores 0.02-0.04 higher than for White patients. This indicates that predicted probabilities are less accurate for minority groups — a calibration-based fairness violation that persists regardless of whether selection rates are equalized. This differential calibration likely reflects underrepresentation in training data or unmeasured heterogeneity within broad racial categories.

### The Consequences of Enforcing Demographic Parity

Table 5 presents the central finding: what happens when demographic parity is enforced through mitigation methods.

**Table 5.**
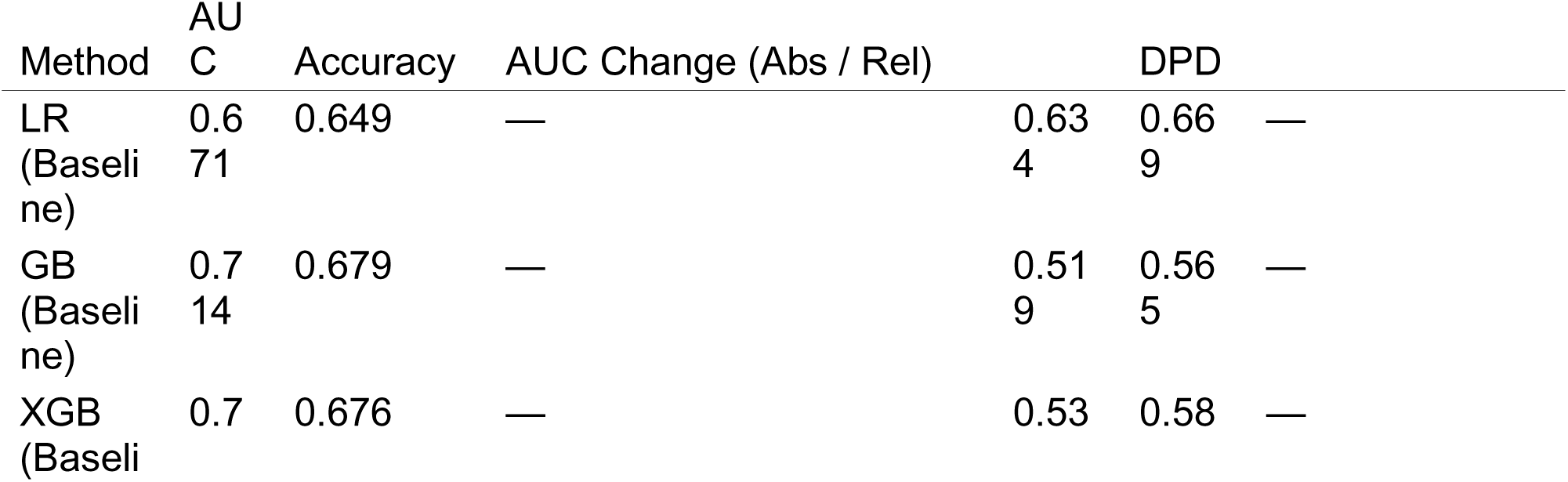
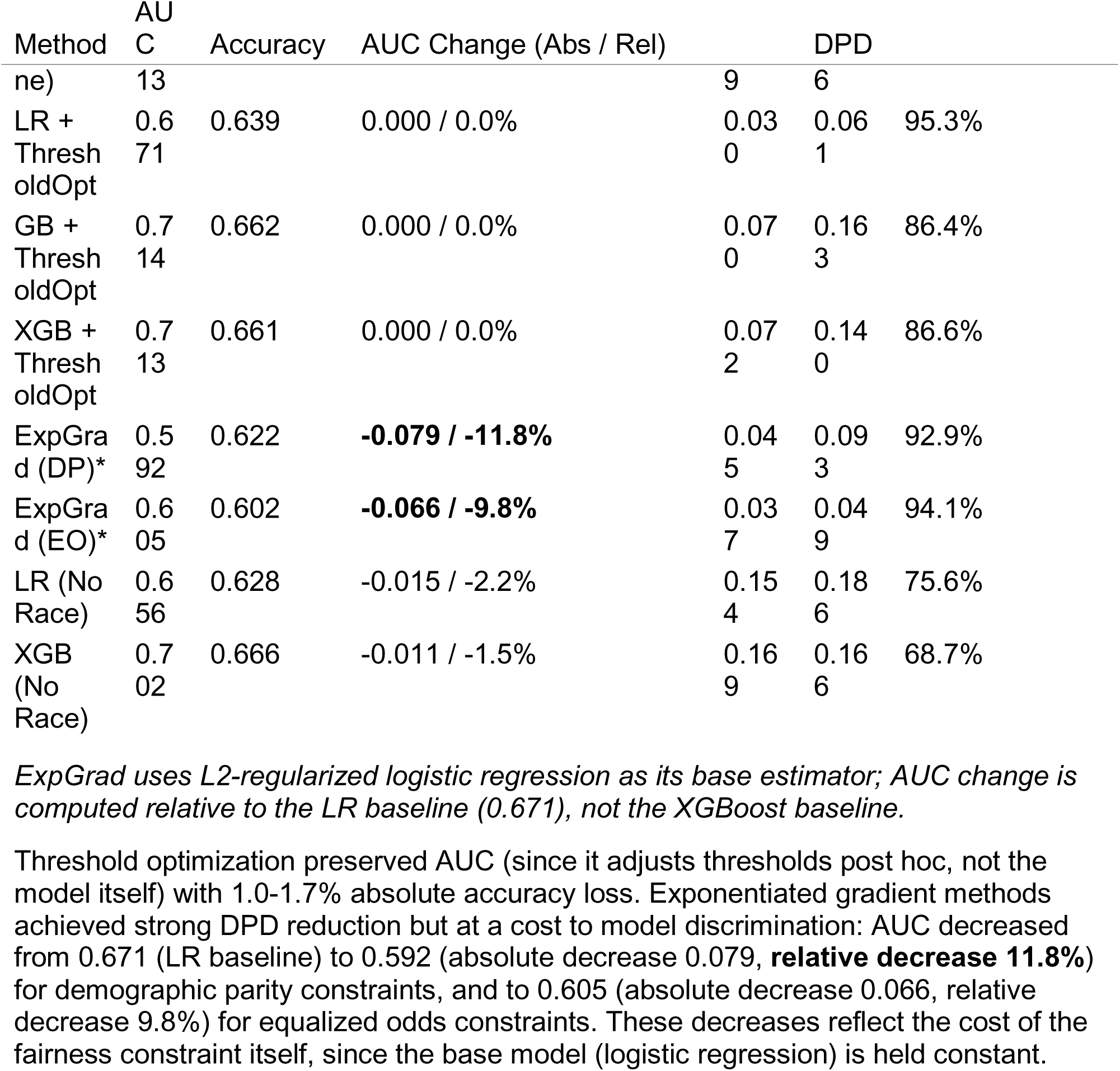
Fairness Mitigation Comparison — Performance and Disparity Trade-offs.

### How Demographic Parity Mitigation Affects High-Burden Groups

Table 6 reveals the clinical consequences of enforcing demographic parity at the group level.

**Table 6.**
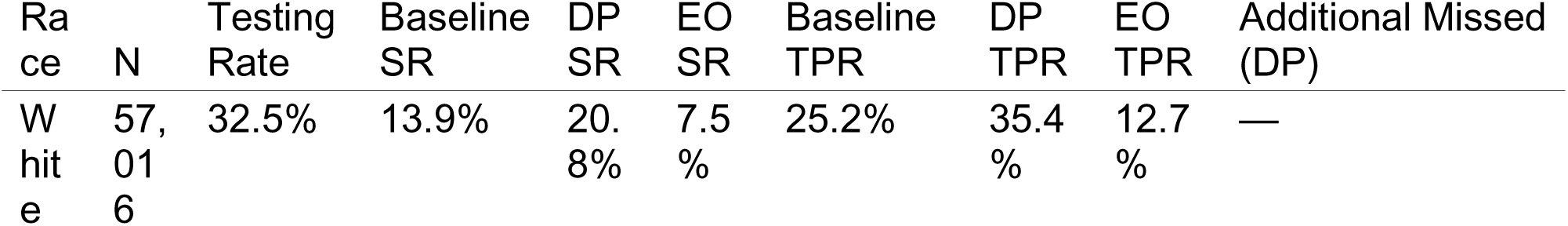
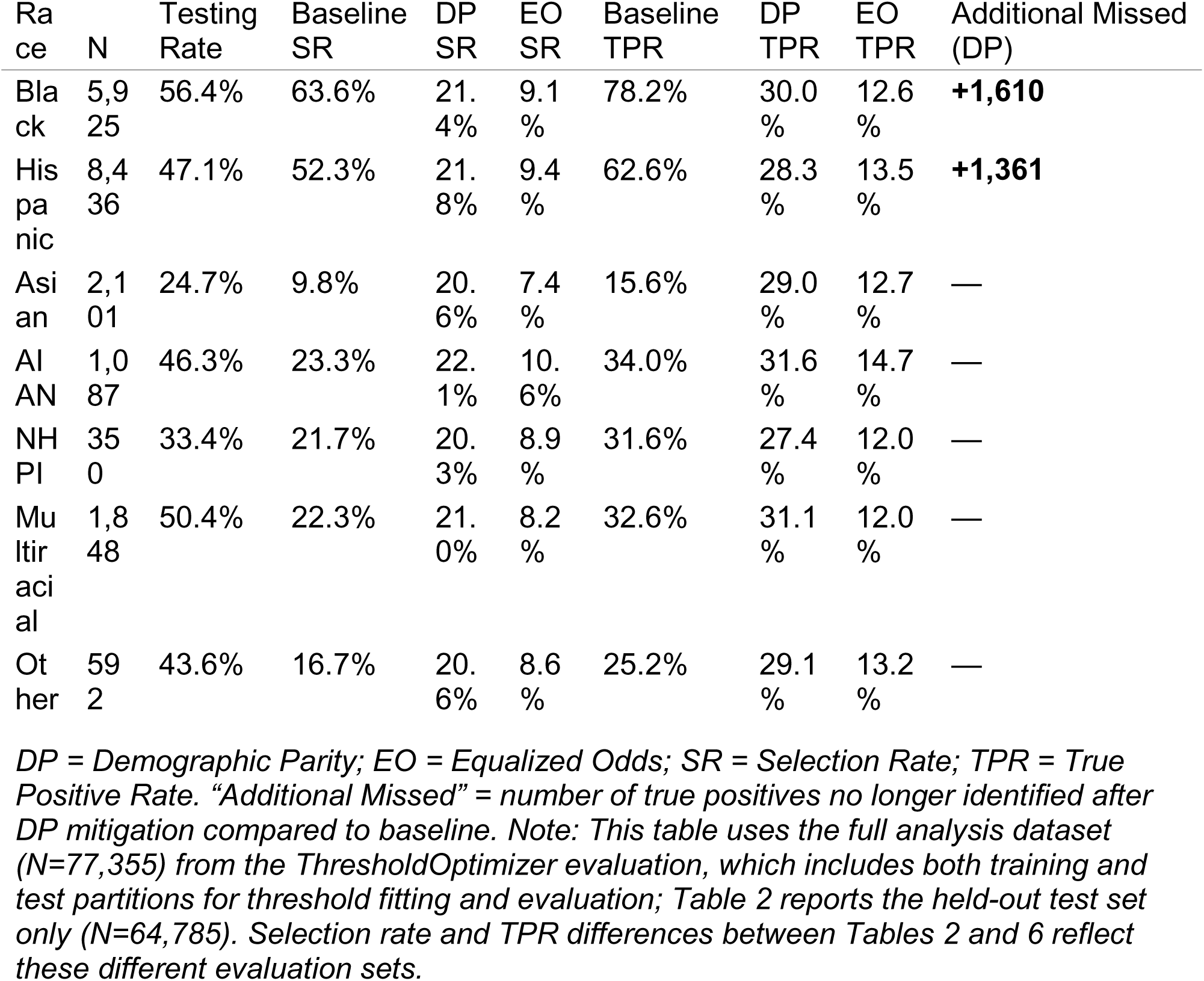
XGBoost + Threshold Optimizer: Performance by Race Before and After Mitigation (Full Analysis Dataset, N=77,355)

| Ra<br>ce | N | Testing<br>Rate | Baseline<br>SR | DP<br>SR | EO<br>SR | Baseline<br>TPR | DP<br>TPR | EO<br>TPR | Additional Missed<br>(DP) |
| --- | --- | --- | --- | --- | --- | --- | --- | --- | --- |
| W<br>hit<br>e | 57,<br>01<br>6 | 32.5% | 13.9% | 20.<br>8% | 7.5<br>% | 25.2% | 35.4<br>% | 12.7<br>% | — |

| Race | N | Testing Rate | Baseline SR | DP SR | EO SR | Baseline TPR | DP TPR | EO TPR | Additional Missed (DP) |
| --- | --- | --- | --- | --- | --- | --- | --- | --- | --- |
| Black | 5,925 | 56.4% | 63.6% | 21.4% | 9.1% | 78.2% | 30.0% | 12.6% | **+1,610** |
| Hispanic | 8,436 | 47.1% | 52.3% | 21.8% | 9.4% | 62.6% | 28.3% | 13.5% | **+1,361** |
| Asian | 2,101 | 24.7% | 9.8% | 20.6% | 7.4% | 15.6% | 29.0% | 12.7% | — |
| AIAN | 1,087 | 46.3% | 23.3% | 22.1% | 10.6% | 34.0% | 31.6% | 14.7% | — |
| NHPI | 350 | 33.4% | 21.7% | 20.3% | 8.9% | 31.6% | 27.4% | 12.0% | — |
| Multiracial | 1,848 | 50.4% | 22.3% | 21.0% | 8.2% | 32.6% | 31.1% | 12.0% | — |
| Other | 592 | 43.6% | 16.7% | 20.6% | 8.6% | 25.2% | 29.1% | 13.2% | — |

*DP = Demographic Parity; EO = Equalized Odds; SR = Selection Rate; TPR = True Positive Rate. “Additional Missed” = number of true positives no longer identified after DP mitigation compared to baseline. Note: This table uses the full analysis dataset (N=77,355) from the ThresholdOptimizer evaluation, which includes both training and test partitions for threshold fitting and evaluation; Table 2 reports the held-out test set only (N=64,785). Selection rate and TPR differences between Tables 2 and 6 reflect these different evaluation sets.*

The pattern is striking. Enforcing demographic parity:

- **Reduced** Black TPR from 78.2% to 30.0% — a 48.2 percentage point (61.6% relative) decrease in sensitivity for the group with the highest HIV testing rate and highest disease burden, resulting in **1,610 additional missed** individuals in the test set alone
- **Reduced** Hispanic TPR from 62.6% to 28.3% — a 34.3 percentage point (54.8% relative) decrease, with **1,361 additional missed**
- **Increased** White TPR from 25.2% to 35.4% and Asian TPR from 15.6% to 29.0%

Equalized odds optimization produced lower overall selection rates and lower TPRs across all groups, reflecting the ThresholdOptimizer’s effort to equalize both TPR and FPR simultaneously — which, given the low baseline FPRs for some groups, required substantial threshold adjustment. The key distinction is conceptual rather than in these particular implementation results: EO as a *criterion* permits differential selection rates proportional to burden, whereas DP as a criterion forces equal selection rates regardless of burden. The ThresholdOptimizer’s EO implementation here equalized at a low TPR level, but alternative implementations (e.g., in-processing EO with epsilon tuning) could target higher overall sensitivity while maintaining TPR parity.

In practical terms, among Black individuals who had been tested for HIV, the baseline model correctly identified 78.2% of them. After demographic parity mitigation, only 30.0% were correctly identified. Extrapolating proportionally to the full BRFSS analysis sample, DP optimization would result in approximately 6,440 additional Black and 5,444 additional Hispanic individuals going unidentified — an approximate but directionally clear estimate of the harm.

**Figure 2.**
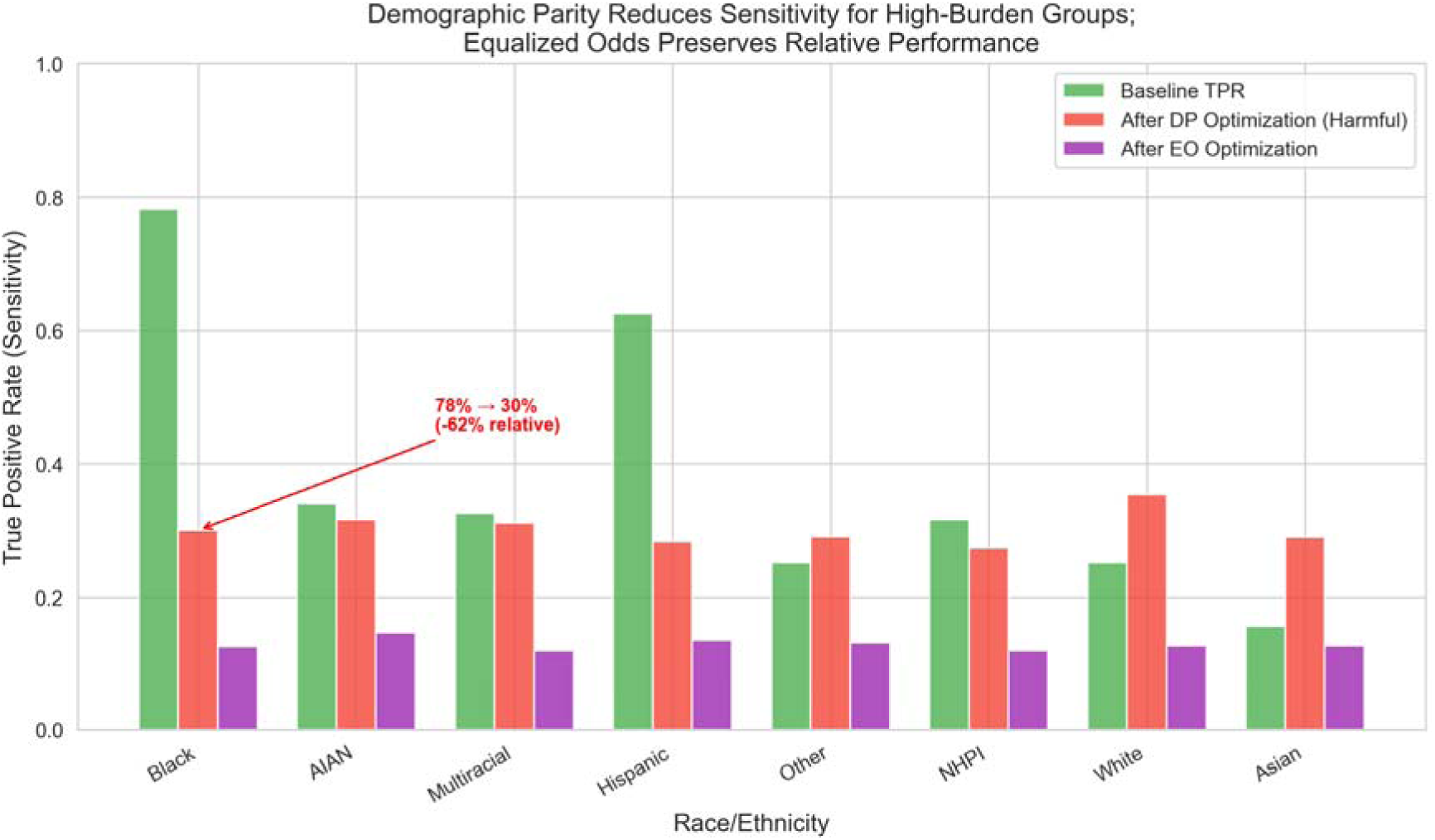
True positive rates (TPR) by race under baseline, demographic parity (DP), and equalized odds (EO) mitigation. DP optimization reduced Black TPR from 78.2% to 30.0% (61.6% relative decrease) and Hispanic TPR from 62.6% to 28.3% (54.8% relative decrease), while increasing White and Asian TPRs — inverting public health screening priorities.

### Intersectional Impact: Race x Sex

Table 7 presents selection rates across intersectional groups, revealing that demographic parity optimization creates particularly severe consequences at the intersection of race and sex.

**Table 7.**
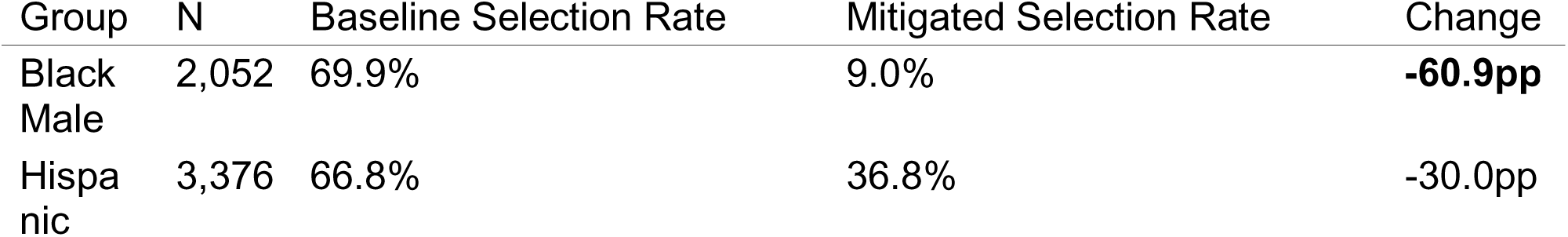
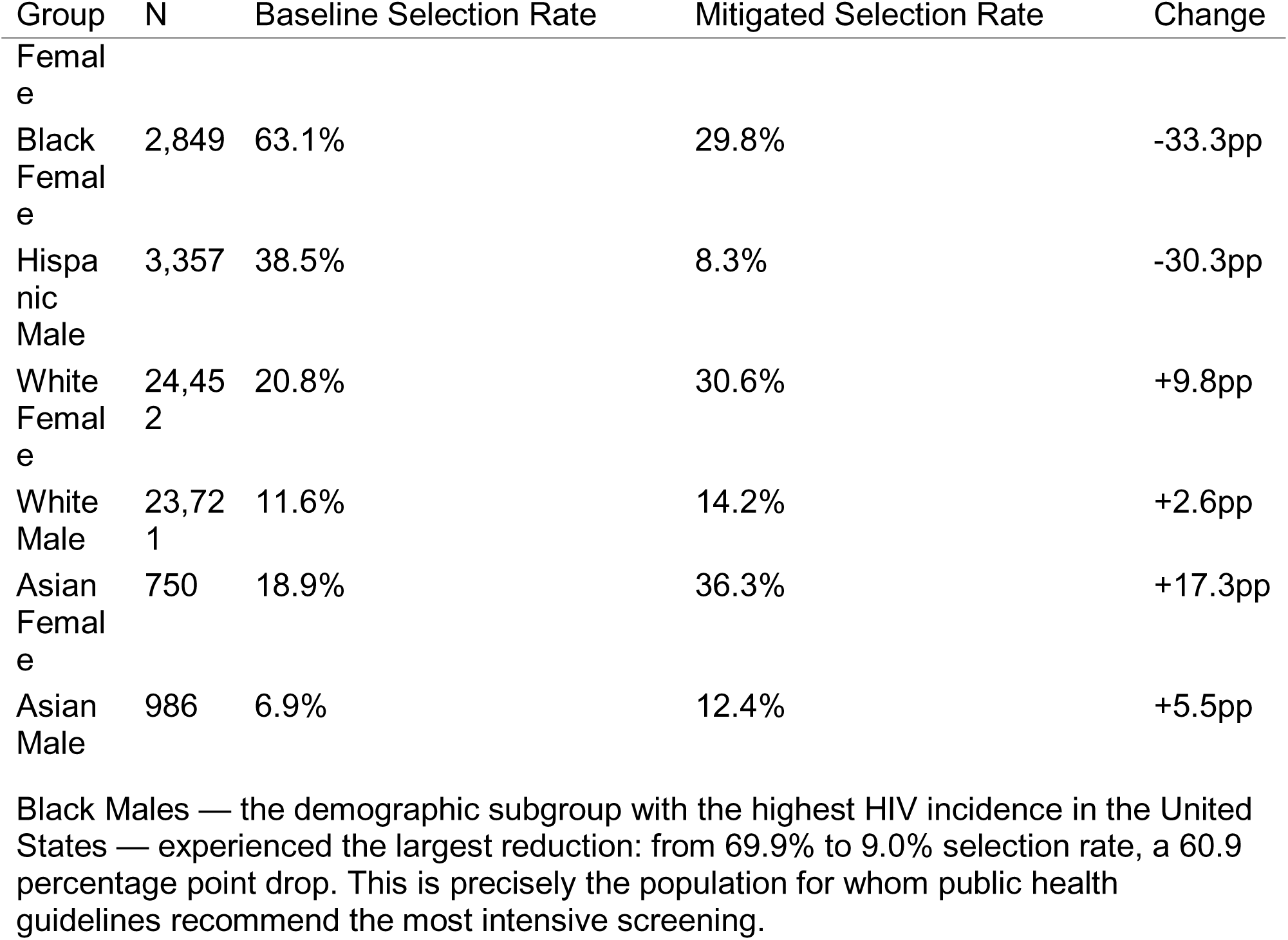
Selection Rates by Race x Sex (Baseline vs. Mitigated)

### Cross-Dimensional Fairness Trade-offs

Optimizing for racial fairness simultaneously degraded sex-based fairness. Sex-based DPD increased from 0.103 (baseline) to 0.176 (after racial DPD optimization) — a 71% worsening (Table 8a). This cross-dimensional transfer of disparity illustrates that single-axis fairness optimization can redistribute rather than eliminate inequity.

To address this, we tested multi-objective optimization approaches that jointly constrain across race and sex (Table 8a).

**Table 8a.**
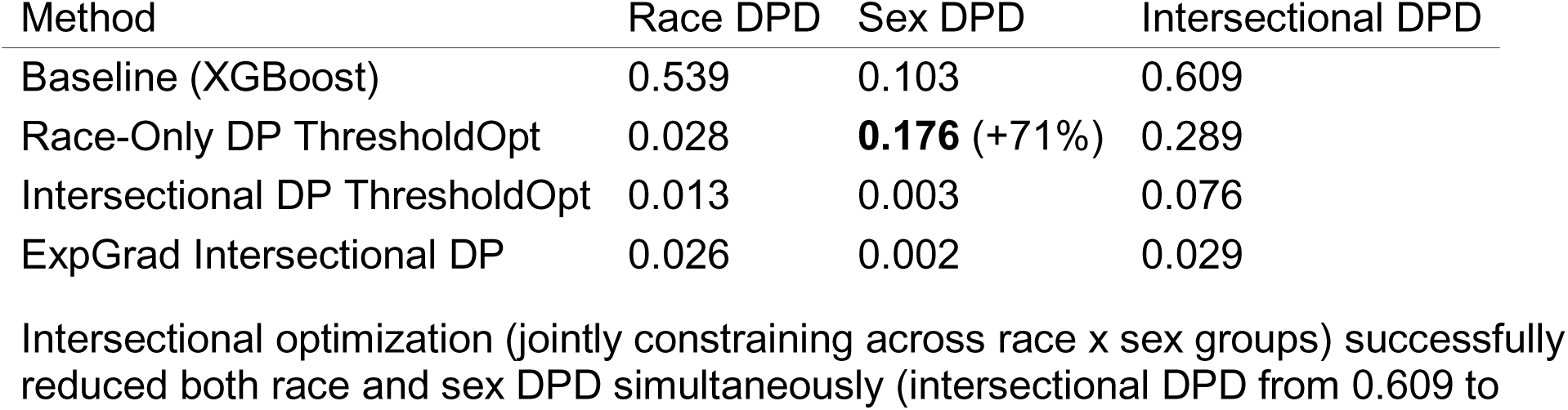
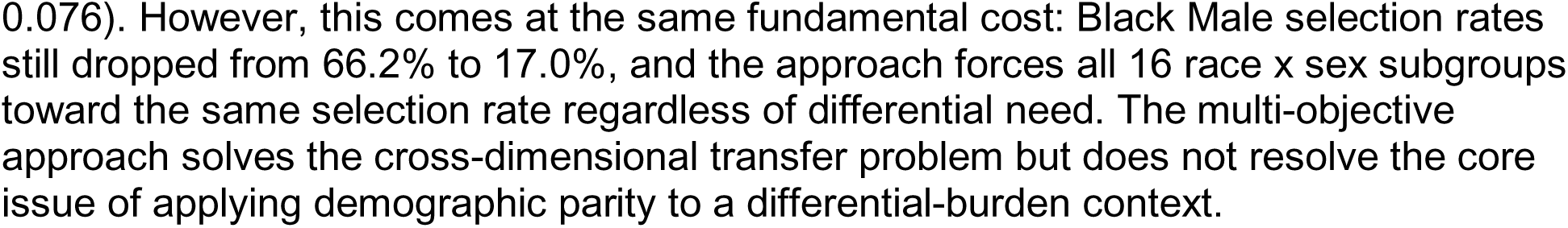
Multi-Objective Fairness Optimization.

**Figure 3.**
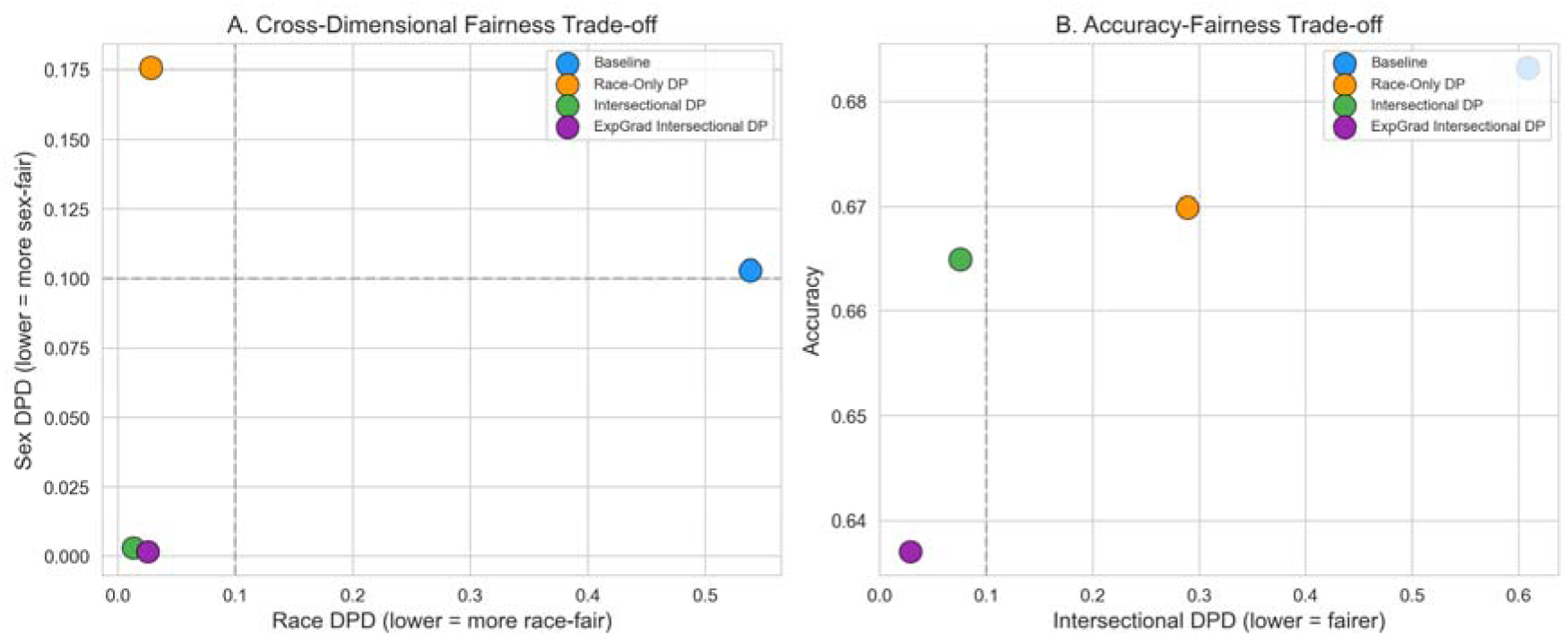
Cross-dimensional fairness trade-offs. (A) Race DPD vs. Sex DPD under different optimization strategies, showing that race-only optimization worsens sex-based disparity by 71%. (B) Intersectional DPD vs. accuracy, illustrating the Pareto frontier of fairness-accuracy trade-offs across mitigation approaches.

### Statistical Testing

**Table 8.** Statistical Tests for Racial Disparities in Selection Rates.

| Comparison | Baseline z | Baseline p | Significant? | Mitigated z | Mitigated p | Significant? |
| --- | --- | --- | --- | --- | --- | --- |
| Black vs White | 81.52 | <0.001 | **Yes** | -2.34 | 0.019 | No* |
| Hispanic vs White | 69.05 | <0.001 | **Yes** | 0.06 | 0.951 | No |
| Asian vs White | -4.66 | <0.001 | **Yes** | 0.15 | 0.880 | No |

\*Not significant after Bonferroni correction ( $\alpha = 0.0167$ )
McNemar's test confirmed that predictions differed significantly before versus after mitigation (chi-squared = 165.89, $p = 5.83 \times 10^{-38}$ ). Chi-square tests showed baseline predictions were strongly dependent on race (chi-squared = 9,450.1, $p < 10^{-300}$ ), while mitigated predictions showed only marginal dependence (chi-squared = 15.3, $p = 0.033$ ).

### Effect Sizes

**Table 9.** Cohen’s h Effect Sizes (Selection Rate Differences vs. White)

| Group | Baseline h | Interpretation | Mitigated h | Interpretation |
| --- | --- | --- | --- | --- |
| Black | 1.065 | **Large** | -0.035 | Negligible |
| Hispanic | 0.794 | **Medium** | 0.001 | Negligible |
| Asian | -0.120 | Small | 0.004 | Negligible |

Effect sizes decreased from large/medium to negligible after mitigation. While this demonstrates that mitigation *can* equalize selection rates, the preceding analysis shows that doing so comes at substantial cost to sensitivity for high-burden groups.

### Sensitivity Analysis: Survey-Weighted Estimates

BRFSS uses a complex survey design with stratification and oversampling. To assess whether our unweighted analyses biased fairness estimates, we recomputed all metrics using BRFSS survey weights (_LLCPWT). Table S1 compares weighted and unweighted results.

**Table S1.** Weighted vs. Unweighted Fairness Metrics (XGBoost)

| Metric | Unweighted | Weighted | Difference |
| --- | --- | --- | --- |
| AUC | 0.716 | 0.703 | -0.013 |
| DPD | 0.539 | 0.574 | +0.036 |
| EOD | 0.625 | 0.666 | +0.040 |

*Note: Minor differences in unweighted AUC (0.716 here vs. 0.713 in Table 1) and EOD (0.625 here vs. 0.586 in Table 1) reflect differences in evaluation methodology: Table 1 reports standard test-set metrics at the default threshold, while this sensitivity analysis recomputes metrics using predicted probabilities with survey-weight-adjusted group calculations on the same test set.*

**Figure 4.**
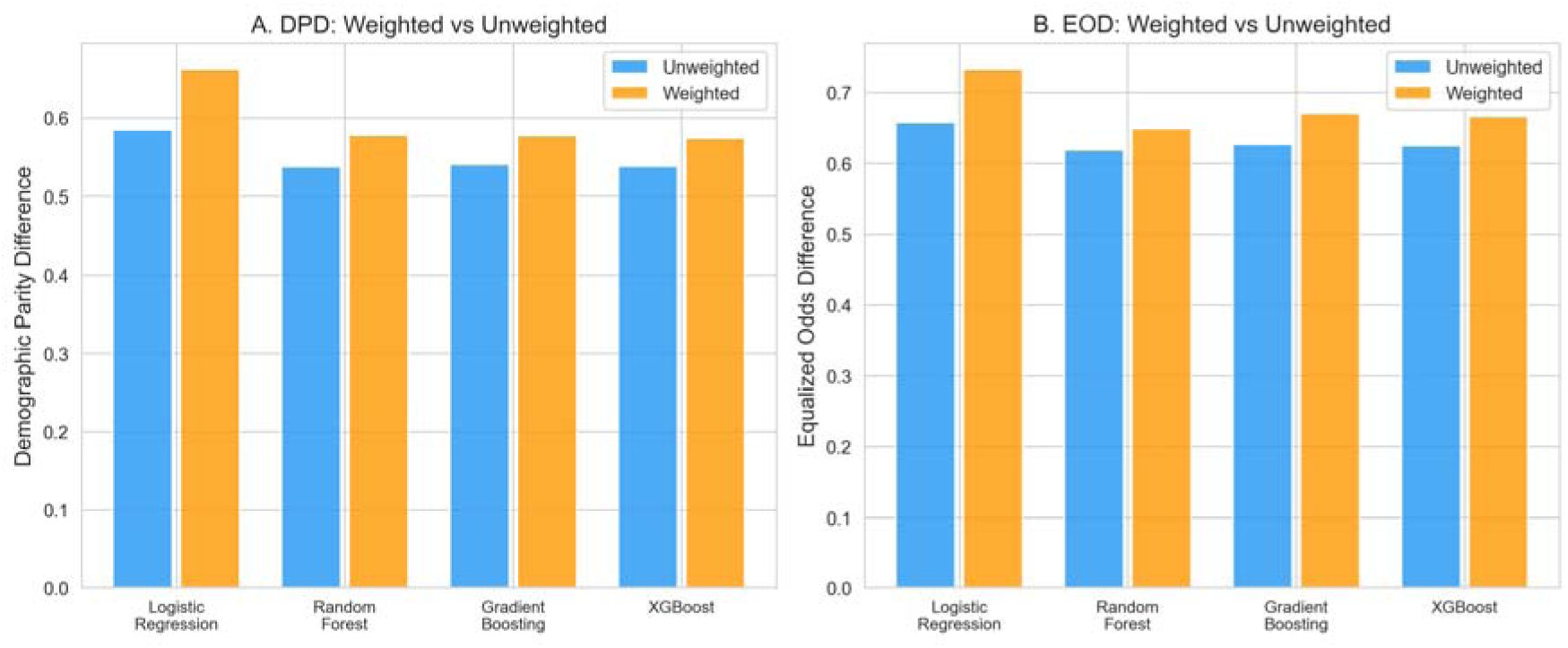
Survey-weighted vs. unweighted fairness metrics across all four classifiers. (A) Demographic Parity Difference (DPD) and (B) Equalized Odds Difference (EOD). Weighted estimates consistently exceed unweighted estimates, indicating that unweighted analyses underestimate the true magnitude of differential prediction.

### Robustness Analyses

**5-Fold Cross-Validation:** Mean DPD reduction was 90.9% +/- 3.1% (95% CI: 84.8%-97.1%) across folds. AUC remained stable at 0.711 +/- 0.002.

**Table 10.** Cross-Validation Results (5 Folds)

| Fold | N_test | Baseline AUC | Baseline DPD | Mitigated DPD | DPD Reduction |
| --- | --- | --- | --- | --- | --- |
| 1 | 64,785 | 0.711 | 0.561 | 0.046 | 91.9% |
| 2 | 64,785 | 0.713 | 0.534 | 0.063 | 88.3% |
| 3 | 64,785 | 0.707 | 0.545 | 0.039 | 92.9% |
| 4 | 64,785 | 0.711 | 0.528 | 0.029 | 94.5% |
| 5 | 64,784 | 0.710 | 0.522 | 0.067 | 87.1% |
| **Mean** | — | **0.711** | **0.538** | **0.049** | **90.9%** |
| **SD** | — | 0.002 | 0.016 | 0.016 | 3.1% |

**Random Seed Stability (10 Seeds):** Mean DPD reduction was 92.4% +/- 2.5%, with range 87.5%-94.9%.

**Bootstrap Confidence Intervals (500 iterations):**

| Model | DPD Point Estimate | 95% CI |
| --- | --- | --- |
| XGB Baseline | 0.539 | [0.518, 0.560] |
| XGB + ThresholdOpt | 0.078 | [0.037, 0.126] |
| ExpGrad (DP) | 0.060 | [0.033, 0.094] |
| XGB No Race | 0.168 | [0.157, 0.181] |

**Sensitivity to Constraint Parameters:** DPD remained stable (0.029-0.054) across epsilon values from 0.001 to 0.2, demonstrating robustness to constraint strength.

**Sensitivity to Base Classifier:** All four base classifiers showed substantial DPD reduction with threshold optimization: LR (96.8%), RF (88.5%), GB (83.9%), XGB (85.2%).

### Coefficient Audit: Denver HIV Risk Score

The Denver Score explicitly assigns differential points by race, with Black patients receiving +7 points and Hispanic patients +3 points regardless of clinical risk factors. A Black male with zero behavioral risk factors scores 15 points (Black +7, male +8) — halfway to the high-risk threshold of 30 — before any clinical assessment. This illustrates explicit race-based stratification in a published HIV risk tool, which may be defensible on epidemiological grounds (reflecting genuine differential incidence) but also embeds demographic profiling into clinical algorithms.

### Clinical Utility Analysis

Decision curve analysis showed that mitigated models maintained positive net benefit across clinically relevant threshold ranges (0.30-0.70).

**Table 11.**
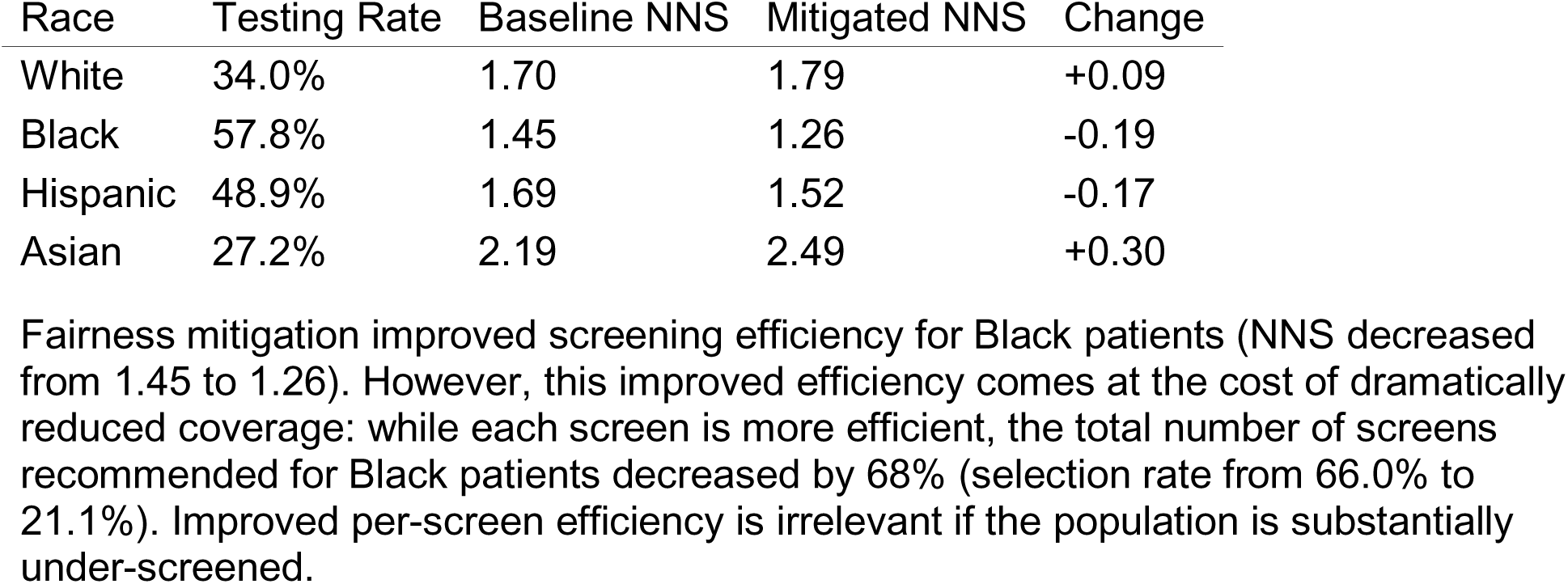
Number Needed to Screen by Race.

## Discussion

### Summary of Findings

The choice of fairness metric has direct clinical consequences when disease burden differs across demographic groups. Using HIV testing prediction as a case study, we show three principal findings:

**First, standard ML classifiers trained on HIV testing data produce differential prediction rates that closely mirror actual differential testing utilization across racial groups.** Selection rates ranged from 12.1% (Asian) to 66.0% (Black), tracking known differences in HIV testing uptake and reflecting the model’s learned representation of existing care patterns. These differential rates exceed conventional fairness thresholds (DPD 0.519-0.634) but do not, by themselves, demonstrate algorithmic bias — they may represent appropriate capture of differential need.

**Second, enforcing demographic parity — equal selection rates regardless of group — actively harms high-burden populations.** Threshold optimization reduced Black TPR from 78.2% to 30.0%, a 61.6% relative decrease in sensitivity, resulting in 1,610 additional missed individuals in the test set. Black Male selection rates dropped from 66.2% to 17.0% under intersectional optimization. In the context of HIV, where Black MSM have the highest incidence of any demographic group, this represents a concrete reduction in screening access for the population most in need. Exponentiated gradient methods incurred additional performance costs, with AUC decreasing from 0.671 (LR baseline) to 0.592 (11.8% relative decrease).

**Third, single-axis fairness optimization transfers disparity across protected dimensions.** Optimizing for racial fairness worsened sex-based disparity by 71% (sex DPD from 0.103 to 0.176), demonstrating that fairness is not a single-dimensional property and that optimization on one axis can create new inequities on another. Multi-objective optimization jointly constraining race and sex reduced intersectional DPD from 0.609 to 0.076, but at the same fundamental cost of suppressing selection rates for high-burden subgroups.

### Why Demographic Parity Is Wrong for HIV Testing

Demographic parity assumes that the base rate of the outcome *should* be equal across groups — that any difference in selection rates constitutes unfairness. This assumption is appropriate in contexts like hiring or lending, where membership in a protected group should be irrelevant to the outcome. It is inappropriate in HIV testing for two reasons:

1. **Unequal disease burden is real.** Black Americans have 8 times the HIV diagnosis rate of White Americans; Hispanic Americans have 4 times the rate (11). These differences are driven by structural factors, not biology, but they are epidemiologically real. A model that recommends more testing for higher-burden populations is allocating resources toward need, not discriminating.
2. **The outcome captures differential utilization, not equal need.** Our model predicts who *has been tested*, not who *should be tested*. Testing rates are higher among Black Americans in part because of decades of targeted public health campaigns in high-burden communities. A model trained on this outcome learns those patterns. Flagging this as “bias” conflates appropriate public health targeting with discrimination. We acknowledge that utilization is an imperfect proxy for need — some utilization differences may reflect access barriers rather than appropriate targeting. However, the directional alignment is clear: groups with higher HIV burden also have higher testing rates, and suppressing those rates through DP optimization moves away from, not toward, appropriate allocation.

The more appropriate fairness criteria for this context are:

- **Equalized odds:** Ensures the model is equally accurate for all groups — among those who were tested, all racial groups should be equally likely to be identified; among those not tested, all groups should have equal false positive rates. This allows higher selection rates for higher-burden groups while demanding equal accuracy.
- **Calibration:** Ensures predicted probabilities are accurate across groups — a 60% prediction means the same thing for a Black patient as for a White patient. Our analysis found calibration differences of 0.02-0.04 in Brier scores, indicating room for improvement.

### The Harm of Naive Fairness Auditing

Our results illustrate a broader risk in healthcare AI fairness: applying fairness frameworks developed for non-clinical contexts (hiring, lending, criminal justice) to clinical settings where differential treatment may be clinically appropriate. The Obermeyer et al. (3) finding — that a health system algorithm disadvantaged Black patients — has rightly heightened awareness of algorithmic bias. But the lesson should not be “all differential prediction is bias.” The lesson should be “differential prediction is concerning and warrants investigation to determine whether it reflects bias or appropriate clinical variation.”

In our case, the investigation reveals that differential prediction reflects differential testing patterns driven by differential disease burden. Enforcing equality of prediction rates in this context does not correct a bias — it creates a new one, by under-recommending screening for high-risk populations.

This finding has direct parallels to recent work on lung cancer screening, where fairness-motivated revisions to USPSTF eligibility criteria were found to potentially reduce screening access for Black smokers who face higher lung cancer risk at lower pack-year thresholds (27,28).

### Mechanisms of Differential Prediction

Our analysis reveals three mechanisms through which models produce differential prediction rates:

**Explicit race coding.** The Denver HIV Risk Score assigns +7 points for Black race and +3 for Hispanic ethnicity. While this reflects epidemiological data, it means demographic profiling is embedded by design.

**Race-correlated features as proxies.** Even in race-blind models, features like insurance status, depression, cost barriers, and geography retain 70% of baseline differential prediction. These features are shaped by the same structural inequities that drive differential HIV burden — poverty, geographic segregation, healthcare access.

Whether capturing these associations constitutes “proxy discrimination” or “appropriate capture of social determinants” depends entirely on how the model’s predictions will be used.

**Differential calibration.** Brier scores were consistently worse for Hispanic patients (0.231-0.251) than White patients (0.199-0.209), indicating that the model’s predictions are less reliable for minority groups. Unlike differential selection rates, this represents a genuine fairness concern: predictions should be equally trustworthy regardless of group membership. Addressing calibration differences — through group-specific calibration, oversampling, or enriched feature sets — would improve model fairness without reducing screening for high-burden populations.

### External Validation: Differential Burden Is Real

To contextualize our findings, we compared model-detected differential prediction rates against real-world clinical outcome data from the Ryan White HIV/AIDS Program (N=372,220 patients), the largest federally funded HIV care program in the United States.

**Table 12.** Ryan White Viral Suppression Rates by Race/Ethnicity (2024)

| Race/Ethnicity | Total Patients | Viral Suppression % | Disparity vs. White |
| --- | --- | --- | --- |
| White | 85,635 | 93.1% | (reference) |
| Asian | 5,548 | 95.8% | +2.7% |
| Hispanic/Latino | 102,377 | 92.0% | -1.1% |
| Black/African American | 172,242 | 88.4% | **-4.7%** |
| AIAN | 1,719 | 88.8% | -4.3% |
| NHPI | 544 | 90.6% | -2.5% |
| Multiple Races | 4,155 | 88.6% | -4.5% |

### Cross-Dimensional Fairness Trade-offs

The finding that racial fairness optimization worsened sex-based fairness (sex DPD increase from 0.103 to 0.176, a 71% degradation) has direct practical implications. Real-world patients belong to multiple protected groups simultaneously. Fairness optimization that considers only one dimension may transfer disparity to another, creating new inequities while resolving old ones. Multi-objective optimization approaches that jointly constrain across multiple protected attributes are needed, but face compounding trade-offs and reduced statistical power for small intersectional subgroups.

### Limitations

Several limitations should be considered.

**Outcome measure.** Our primary outcome was lifetime HIV testing history rather than clinical HIV risk, recent testing, or HIV status. This limits our ability to assess whether differential prediction reflects appropriate risk stratification versus bias. Datasets with HIV diagnosis outcomes (e.g., electronic health records, claims data) would enable need-conditioned fairness analysis using metrics such as equalized odds with clinical outcomes as the ground truth. We note that BRFSS includes a variable for most recent HIV test date (HIVTSTD3), which could be used to derive more proximal measures.

**Survey weights.** BRFSS uses a complex survey design with stratification, clustering, and oversampling. Our primary analyses treat the data as an unweighted sample.

Sensitivity analysis using BRFSS survey weights (_LLCPWT) showed that weighted estimates yielded slightly larger disparities (DPD 6.7% higher, EOD 6.4% higher), indicating that unweighted analyses underestimate rather than overestimate disparities. All qualitative conclusions were unchanged. However, our weighted analysis used only the final sampling weight and did not incorporate the full complex survey design (stratification and PSU variables) for variance estimation, which may affect confidence interval widths.

**Missing behavioral variables.** BRFSS does not include behavioral risk variables (MSM status, injection drug use, condom use) that are central to clinical HIV risk assessment. Our models therefore rely on demographic and socioeconomic predictors that are proxies for — rather than direct measures of — HIV risk. This limits the clinical applicability of the models themselves, though it does not undermine our central methodological finding about fairness metric selection.

**Limited fairness criteria evaluated.** We focused on demographic parity, equalized odds, and calibration. Other criteria — equal opportunity (TPR parity only), needs-proportional allocation, or conditional use accuracy equality — may be more appropriate for specific deployment contexts and should be examined in future work.

**Intersectional limitations.** Our intersectional analysis examined race x sex (16 groups), but more comprehensive approaches examining interactions with age, geography, insurance status, and sexual orientation would provide additional insight.

**Sampling limitations.** BRFSS uses telephone-based sampling that may underrepresent populations without stable housing or phone access — precisely those at highest HIV risk.

### Future Directions

This work motivates several lines of inquiry:

**Need-conditioned fairness analysis.** The most important next step is replicating this analysis using datasets with HIV diagnosis outcomes (EHR or claims data), enabling fairness assessment conditioned on clinical need rather than testing utilization. This would allow direct evaluation of whether models over-or under-recommend testing relative to actual risk.

**Auditing deployed models.** Published HIV screening models — including the Denver Score and recently developed EHR-based PrEP candidacy algorithms (15,16) — should undergo fairness audits using need-appropriate metrics. May et al. (29) recently developed a generalizable HIV risk prediction pipeline across EHR systems that would be a suitable candidate for such an audit.

**Stakeholder-driven metric selection.** The choice between fairness criteria (demographic parity vs. equalized odds vs. calibration vs. needs-based allocation) involves value judgments that should be informed by clinicians, patients, community advocates, and ethicists — not determined by algorithm developers alone. Participatory approaches to fairness metric selection in HIV prevention are needed.

**Multi-dimensional fairness optimization.** Methods that jointly optimize across multiple protected attributes (race, sex, age, geography) while respecting clinical priorities warrant development and evaluation.

## Conclusions

In healthcare contexts where disease burden differs across demographic groups, the choice of fairness metric is not a technical detail — it is a clinical decision with direct consequences for patient care. We demonstrate that enforcing demographic parity in HIV testing prediction reduces screening sensitivity for Black Americans by 61.6%, resulting in 1,610 additional missed individuals in our test set alone, effectively de-prioritizing the population with the highest HIV burden. Multi-objective optimization across race and sex jointly reduced intersectional disparities but imposed the same fundamental cost. Race-blind models retain 70% of differential prediction through correlated social determinants, but this differential prediction may reflect appropriate capture of need-associated factors rather than bias.

We recommend that fairness audits in healthcare: (1) use need-appropriate metrics — equalized odds and calibration — rather than defaulting to demographic parity; (2) quantify the clinical consequences of mitigation, not just the statistical properties; (3) involve stakeholder deliberation on fairness criteria appropriate to the clinical context; and (4) explicitly examine cross-dimensional trade-offs when optimizing for fairness on a single protected attribute. Algorithmic fairness in healthcare requires clinical judgment, not just mathematical optimization.

## Data Availability

The BRFSS 2024 dataset is publicly available from the CDC at https://www.cdc.gov/brfss/. NHANES data are available at https://www.cdc.gov/nchs/nhanes/. Ryan White aggregate data are available from HRSA at https://ryanwhite.hrsa.gov/data/reports. AIDSVu data are available at https://aidsvu.org/. CDC AtlasPlus data are available at https://www.cdc.gov/nchhstp/atlas/. Analysis code is available at https://github.com/hayden-farquhar/HIV-testing-fairness.

## Data Availability

All data used in this study are publicly available. The BRFSS 2024 dataset is available from the CDC at https://www.cdc.gov/brfss/. Ryan White HIV/AIDS Program data are available from HRSA at https://ryanwhite.hrsa.gov/data/reports. AIDSVu county-level data are available at https://aidsvu.org/. Analysis code is available at https://github.com/hayden-farquhar/HIV-testing-fairness

## Acknowledgments

I gratefully acknowledge the data sources that made this research possible:

- The **Centers for Disease Control and Prevention (CDC)** for the Behavioral Risk Factor Surveillance System (BRFSS) 2024 dataset and the participants who contributed their health information through this survey
- The **National Center for Health Statistics** for the National Health and Nutrition Examination Survey (NHANES) data
- The **Health Resources and Services Administration (HRSA)** for the Ryan White HIV/AIDS Program Annual Client-Level Data, and the patients and providers participating in the Ryan White Program
- **AIDSVu** (Emory University Rollins School of Public Health) for county-level HIV prevalence and care continuum data
- **The** CDC National Center for HIV, Viral Hepatitis, STD, and TB Prevention for the AtlasPlus HIV surveillance data

I also acknowledge the developers of the Fairlearn library (Microsoft Research), whose open-source tools enabled the fairness analyses presented in this work.

## Ethics Statement

This study used publicly available, de-identified survey data (BRFSS, NHANES) and aggregate program data (Ryan White, CDC AtlasPlus, AIDSVu). No individual-level identifiable data were accessed. The study was exempt from institutional review board review.

## Author Contributions

HF conceived the study, designed the analysis, wrote the code, performed all analyses, and wrote the manuscript.

## Use of AI Tools

AI writing assistance (Claude, Anthropic) was used for manuscript drafting and editing. All analyses, interpretation of results, and scientific conclusions are the sole responsibility of the author. All AI-assisted text was reviewed and revised by the author.

## References

1. Chen IY, et al. Can AI help reduce disparities in general medical and mental health care? AMA J Ethics. 2019;21:E167–179.

2. Mehrabi N, et al. A survey on bias and fairness in machine learning. ACM Comput Surv. 2021;54:1–35.

3. Obermeyer Z, et al. Dissecting racial bias in an algorithm used to manage the health of populations. Science. 2019;366:447–453.

4. Adamson AS, Smith A. Machine learning and health care disparities in dermatology. JAMA Dermatol. 2018;154:1247–1248.

5. Wong A, et al. External validation of a widely implemented proprietary sepsis prediction model in hospitalized patients. JAMA Intern Med. 2021;181:1065–1070.

6. Vyas DA, et al. Hidden in plain sight — Reconsidering the use of race correction in clinical algorithms. N Engl J Med. 2020;383:874–882.

7. Chouldechova A. Fair prediction with disparate impact. Big Data. 2017;5:153–163.

8. Corbett-Davies S, Goel S. The measure and mismeasure of fairness: a critical review of fair machine learning. J Mach Learn Res. 2023;24(312):1–117.

9. Kleinberg J, et al. Inherent trade-offs in the fair determination of risk scores. Innov Theor Comput Sci. 2017;43:1–23.

10. Hardt M, Price E, Srebro N. Equality of opportunity in supervised learning. arXiv:1610.02413. 2016.

11. Centers for Disease Control and Prevention. HIV Surveillance Report, 2021. 2023.

12. Maulsby C, et al. HIV among Black men who have sex with men in the United States. AIDS Behav. 2014;18:10–25.

13. Pellowski JA, et al. A pandemic of the poor: Social disadvantage and the U.S. HIV epidemic. Am Psychol. 2013;68:197–209.

14. Calabrese SK, et al. The impact of patient race on clinical decisions related to prescribing HIV pre-exposure prophylaxis (PrEP). AIDS Behav. 2014;18:226–240.

15. Marcus JL, et al. Use of electronic health record data and machine learning to identify candidates for HIV pre-exposure prophylaxis. Lancet HIV. 2019;6:e688–e695.

16. Krakower DS, et al. Development and validation of an automated HIV prediction algorithm. Lancet HIV. 2019;6:e696–e704.

17. Petersen ML, et al. Super learner analysis of electronic adherence data improves viral prediction. J Acquir Immune Defic Syndr. 2015;69:109–118.

18. Crawford TN. Poor retention in care one-year after viral suppression. AIDS Care. 2014;26:1393–1399.

19. Dombrowski JC, et al. Population-based metrics for the timing of HIV diagnosis, engagement in HIV care, and virologic suppression. AIDS. 2012;26:77–86.

20. Mugavero MJ, et al. Measuring retention in HIV care: the elusive gold standard. J Acquir Immune Defic Syndr. 2012;61:574–580.

21. Juusola JL, Brandeau ML. HIV treatment and prevention: a simple model to determine optimal investment. Med Decis Making. 2016;36:391–409.

22. Ridgway JP, et al. Which patients in the emergency department should receive preexposure prophylaxis? AIDS Patient Care STDs. 2018;32:202–207.

23. Haukoos JS, et al. Derivation and validation of the Denver Human Immunodeficiency Virus (HIV) Risk Score for targeted HIV screening. Am J Epidemiol. 2012;175:838–846.

24. Bird S, et al. Fairlearn: A toolkit for assessing and improving fairness in AI. Microsoft Research. 2020.

25. Agarwal A, et al. A reductions approach to fair classification. ICML. 2018;35:60–69.

26. Vickers AJ, Elkin EB. Decision curve analysis. Med Decis Making. 2006;26:565–574.

27. Aldrich MC, et al. Evaluation of USPSTF Lung Cancer Screening Guidelines Among African American Adult Smokers. JAMA Oncol. 2019;5(9):1318.

28. Manful A, Mercaldo S, Blume JD, Aldrich MC. Addressing algorithmic bias in lung cancer screening eligibility. JNCI. 2025. doi:10.1093/jnci/djaf298.

29. May S, Giordano TP, Gottlieb A. Generalizable pipeline for constructing HIV risk prediction models across EHR systems. JAMIA. 2024;31(3):666–673.

30. Ryan White HIV/AIDS Program. Annual Client-Level Data Report. HRSA; 2024.

31. AIDSVu. Local HIV data. Emory University Rollins School of Public Health. 2024.

