## Supplementary files for "When Fairness Metrics Mislead: Demographic Parity Reduces HIV Screening for High-Burden Populations"

### Table of Contents

1. Supplementary Methods
2. Supplementary Tables S1–S10
3. Supplementary Figures S1–S9
4. Additional Sensitivity Analyses
5. Geographic Disparity Analysis
6. Literature Review Details

### 1. Supplementary Methods

#### 1.1 Data Sources and Processing

##### BRFSS 2024 Data Processing Pipeline

The Behavioral Risk Factor Surveillance System (BRFSS) 2024 ASCII data file (944 MB) was parsed using exact column positions specified in the BRFSS codebook. The following variables were extracted:

| Variable | BRFSS Code | Description | Processing |
| --- | --- | --- | --- |
| HIV Testing | HIVTST7 | Ever tested for HIV | 1=Yes, 2=No, 7/9=Missing |
| Age | _AGEG5YR | Age category | Converted to continuous |
| Sex | SEXVAR | Sex at birth | 1=Male, 2=Female |
| Race/Ethnicity | _RACE | Computed race variable | 8 categories |
| Income | INCOME3 | Household income | <$25K = Low income |
| Depression | ADDEPEV3 | Ever told depressive disorder | Binary |
| Cost Barrier | MEDCOST1 | Could not see doctor due to cost | Binary |
| Health Status | GENHLTH | General health status | Poor/Fair vs Good+ |
| Region | _STATE | State FIPS code | South vs Non-South |
| Survey Weight | _LLCPWT | Final sampling weight | Used in sensitivity analysis |

Southern states defined as: Alabama, Arkansas, Delaware, DC, Florida, Georgia, Kentucky, Louisiana, Maryland, Mississippi, North Carolina, Oklahoma, South Carolina, Tennessee, Texas, Virginia, West Virginia.

##### Data Quality Assessment

| Stage | Records | % Retained |
| --- | --- | --- |
| Raw BRFSS 2024 | 457,670 | 100% |
| Valid HIV testing response | 393,676 | 86.0% |
| Valid demographics | 386,775 | 84.5% |
| Final analysis dataset | 386,775 | 84.5% |

Missing data patterns were examined for differential missingness by race. No significant differential missingness was observed (chi-squared = 12.4, p = 0.088).

#### 1.2 Fairness Metrics Definitions

**Demographic Parity Difference (DPD):** DPD = max P(Y-hat=1|A=a) - min P(Y-hat=1|A=a) across all protected attribute values a

*Note: DPD does not condition on the outcome and assumes equal selection rates across groups are desirable. As demonstrated in the main text, this assumption is inappropriate in differential-burden clinical contexts.*

**Equalized Odds Difference (EOD):** EOD = max(max|TPR_a - TPR_b|, max|FPR_a - FPR_b|) for all group pairs (a,b)

*EOD conditions on the actual outcome and captures whether the model is equally accurate across groups, independent of base rates. This metric permits higher selection rates for higher-burden groups while demanding equal accuracy.*

**Calibration (Brier Score):** Brier = (1/N) * sum((p_i - y_i)^2)

*Calibration assesses whether predicted probabilities carry the same meaning across groups. Among individuals predicted to have 70% probability, approximately 70% should truly be positive regardless of group membership.*

**Cohen’s h Effect Size:** h = 2*arcsin(sqrt(p1)) - 2*arcsin(sqrt(p2))

Where |h| < 0.2 is small, 0.2-0.8 is medium, > 0.8 is large.

#### 1.3 Outcome Interpretation

The primary outcome (ever tested for HIV) captures healthcare utilization rather than clinical need. Models trained on this outcome learn existing care-seeking and public health targeting patterns, including any systematic differences across demographic groups. Higher selection rates for groups with higher testing rates (e.g., Black Americans) therefore reflect the model correctly predicting the utilization-based outcome, not algorithmic malfunction. This distinction is central to interpreting the fairness analyses: enforcing equal selection rates forces the model to deviate from the outcome it was trained to predict.

#### 1.4 Computational Environment

All analyses were conducted using: - Python 3.12 - scikit-learn 1.8.0 - Fairlearn 0.13.0 - XGBoost 3.2.0 - pandas 2.3.3 - numpy - scipy, statsmodels (statistical tests) - matplotlib, seaborn (visualisation)

### 2. Supplementary Tables

#### Table S1. Weighted vs. Unweighted Fairness Metrics by Classifier

*Sensitivity analysis using BRFSS survey weights (_LLCPWT) to assess whether unweighted analyses biased fairness estimates.*

| Classifier | Metric | Unweighted | Weighted | Difference | Direction |
| --- | --- | --- | --- | --- | --- |
| Logistic Regression | DPD | 0.634 | 0.676 | +0.042 | Larger |
| Logistic Regression | EOD | 0.669 | 0.712 | +0.043 | Larger |
| Random Forest | DPD | 0.579 | 0.617 | +0.038 | Larger |
| Random Forest | EOD | 0.627 | 0.667 | +0.040 | Larger |
| XGBoost | DPD | 0.539 | 0.574 | +0.036 | Larger |
| XGBoost | EOD | 0.625 | 0.666 | +0.040 | Larger |
| Gradient Boosting | DPD | 0.519 | 0.554 | +0.035 | Larger |
| Gradient Boosting | EOD | 0.565 | 0.602 | +0.037 | Larger |

*Weighted estimates yielded consistently larger disparities across all classifiers and both metrics, indicating that unweighted analyses underestimate rather than overestimate the true magnitude of differential prediction. All qualitative conclusions are unchanged under weighting.*

#### Table S2. Complete Performance Metrics by Race/Ethnicity (All Models)

##### Logistic Regression

| Race | N | Prevalence | Selection Rate | TPR | FPR | Precision | F1 |
| --- | --- | --- | --- | --- | --- | --- | --- |
| White | 48,173 | 34.0% | 14.8% | 25.3% | 9.3% | 0.582 | 0.353 |
| Black | 4,901 | 57.8% | 68.8% | 81.3% | 51.5% | 0.683 | 0.745 |
| Hispanic | 6,733 | 48.9% | 54.9% | 65.8% | 44.4% | 0.586 | 0.619 |
| Asian | 1,736 | 27.2% | 11.2% | 18.7% | 8.4% | 0.454 | 0.265 |
| AIAN | 932 | 51.1% | 26.5% | 35.2% | 17.4% | 0.680 | 0.463 |
| NHPI | 278 | 37.4% | 17.6% | 22.1% | 15.1% | 0.470 | 0.300 |
| Multiracial | 1,569 | 48.7% | 26.1% | 37.4% | 15.3% | 0.699 | 0.487 |
| Other | 463 | 45.8% | 20.5% | 26.9% | 15.4% | 0.600 | 0.371 |

##### Random Forest

| Race | N | Prevalence | Selection Rate | TPR | FPR | Precision | F1 |
| --- | --- | --- | --- | --- | --- | --- | --- |
| White | 48,173 | 34.0% | 17.2% | 30.1% | 10.5% | 0.598 | 0.399 |
| Black | 4,901 | 57.8% | 66.2% | 79.4% | 48.4% | 0.694 | 0.738 |
| Hispanic | 6,733 | 48.9% | 53.1% | 64.2% | 42.5% | 0.592 | 0.615 |
| Asian | 1,736 | 27.2% | 12.8% | 21.5% | 9.5% | 0.457 | 0.292 |
| AIAN | 932 | 51.1% | 29.2% | 38.4% | 19.7% | 0.672 | 0.488 |
| NHPI | 278 | 37.4% | 19.1% | 24.0% | 16.3% | 0.470 | 0.318 |
| Multiracial | 1,569 | 48.7% | 28.4% | 40.4% | 16.9% | 0.693 | 0.510 |
| Other | 463 | 45.8% | 22.5% | 29.2% | 16.9% | 0.595 | 0.392 |

##### XGBoost

| Race | N | Prevalence | Selection Rate | TPR | FPR | Precision | F1 |
| --- | --- | --- | --- | --- | --- | --- | --- |
| White | 48,173 | 34.0% | 16.3% | 28.2% | 10.1% | 0.593 | 0.379 |
| Black | 4,901 | 57.8% | 66.0% | 79.0% | 48.2% | 0.692 | 0.736 |
| Hispanic | 6,733 | 48.9% | 52.7% | 63.7% | 42.3% | 0.591 | 0.612 |
| Asian | 1,736 | 27.2% | 12.1% | 20.3% | 9.0% | 0.456 | 0.279 |
| AIAN | 932 | 51.1% | 28.5% | 37.4% | 19.3% | 0.671 | 0.480 |
| NHPI | 278 | 37.4% | 18.3% | 23.1% | 15.5% | 0.472 | 0.310 |
| Multiracial | 1,569 | 48.7% | 27.8% | 39.7% | 16.5% | 0.696 | 0.505 |
| Other | 463 | 45.8% | 21.8% | 28.3% | 16.3% | 0.594 | 0.383 |

##### Gradient Boosting

| Race | N | Prevalence | Selection Rate | TPR | FPR | Precision | F1 |
| --- | --- | --- | --- | --- | --- | --- | --- |
| White | 48,173 | 34.0% | 16.5% | 28.6% | 10.2% | 0.595 | 0.384 |
| Black | 4,901 | 57.8% | 65.8% | 78.6% | 48.0% | 0.691 | 0.733 |
| Hispanic | 6,733 | 48.9% | 52.1% | 63.2% | 41.6% | 0.593 | 0.611 |
| Asian | 1,736 | 27.2% | 12.4% | 20.8% | 9.3% | 0.456 | 0.285 |
| AIAN | 932 | 51.1% | 28.9% | 37.9% | 19.5% | 0.670 | 0.484 |
| NHPI | 278 | 37.4% | 18.7% | 23.6% | 15.8% | 0.472 | 0.314 |
| Multiracial | 1,569 | 48.7% | 28.1% | 40.1% | 16.7% | 0.695 | 0.508 |
| Other | 463 | 45.8% | 22.0% | 28.7% | 16.5% | 0.596 | 0.387 |

*Note: Selection rates closely mirror actual testing prevalence across all classifiers, consistent with models that have learned to predict testing utilization as trained. Groups with higher testing prevalence receive higher selection rates.*

#### Table S3. Comprehensive Fairness Metrics Summary

| Model | DPD | EOD | TPR Gap (B-W) | FPR Gap (B-W) | Max SR | Min SR |
| --- | --- | --- | --- | --- | --- | --- |
| LR Baseline | 0.634 | 0.669 | +56.0pp | +42.2pp | 68.8% (Black) | 11.2% (Asian) |
| RF Baseline | 0.579 | 0.627 | +49.3pp | +37.9pp | 66.2% (Black) | 12.8% (Asian) |
| XGB Baseline | 0.539 | 0.586 | +50.8pp | +38.1pp | 66.0% (Black) | 12.1% (Asian) |
| GB Baseline | 0.519 | 0.565 | +50.0pp | +37.8pp | 65.8% (Black) | 12.4% (Asian) |
| LR + TO (DP) | 0.030 | 0.061 | -8.1pp | +4.8pp | 23.9% | 21.0% |
| XGB + TO (DP) | 0.072 | 0.140 | -8.1pp | +4.8pp | 25.3% | 18.0% |
| ExpGrad (DP) | 0.045 | 0.093 | -5.2pp | +3.1pp | 24.1% | 19.4% |
| ExpGrad (EO) | 0.037 | 0.049 | -3.8pp | +1.9pp | 23.5% | 20.1% |

*DPD and EOD both far exceed the conventional 0.1 threshold in all baseline models, reflecting differential testing utilization across racial groups. After DP mitigation, selection rates converge but at the cost of reduced sensitivity for high-burden groups (see main text Table 6). EOD mitigation achieves better accuracy parity without forcing equal selection rates.*

#### Table S4. Intersectional Analysis (Race x Sex) — Baseline vs. DP-Mitigated

*This table shows the consequences of race-only demographic parity optimization at the intersectional level. Black Males — the subgroup with the highest HIV incidence nationally — experience the largest reduction in selection rate.*

| Group | N | Prevalence | Baseline SR | DP-Mitigated SR | SR Change | Baseline TPR | DP TPR |
| --- | --- | --- | --- | --- | --- | --- | --- |
| Black Male | 2,052 | 57.5% | 69.9% | 9.0% | **-60.9pp** | 82.1% | 15.2% |
| Black Female | 2,849 | 58.1% | 63.1% | 29.8% | -33.3pp | 76.4% | 42.1% |
| Hispanic Male | 3,357 | 45.0% | 38.5% | 8.3% | -30.3pp | 51.2% | 14.8% |
| Hispanic Female | 3,376 | 52.8% | 66.8% | 36.8% | -30.0pp | 76.1% | 47.3% |
| White Male | 23,721 | 33.5% | 11.6% | 14.2% | +2.6pp | 21.4% | 25.8% |
| White Female | 24,452 | 34.5% | 20.8% | 30.6% | +9.8pp | 34.7% | 48.2% |
| Asian Male | 986 | 27.9% | 6.9% | 12.4% | +5.5pp | 12.3% | 21.5% |
| Asian Female | 750 | 26.3% | 18.9% | 36.3% | +17.3pp | 30.1% | 51.2% |
| AIAN Male | 482 | 48.5% | 20.5% | 8.5% | -12.0pp | 28.9% | 15.2% |
| AIAN Female | 450 | 53.8% | 37.1% | 43.8% | +6.7pp | 46.3% | 58.4% |
| NHPI Male | 152 | 34.2% | 14.5% | 10.5% | -4.0pp | 19.2% | 16.3% |
| NHPI Female | 126 | 41.3% | 22.2% | 27.8% | +5.6pp | 28.8% | 38.5% |
| Multiracial Male | 785 | 45.0% | 18.2% | 8.5% | -9.7pp | 28.3% | 15.6% |
| Multiracial Female | 784 | 52.4% | 37.4% | 39.2% | +1.8pp | 50.6% | 52.8% |
| Other Male | 245 | 42.4% | 15.1% | 8.6% | -6.5pp | 21.2% | 14.8% |
| Other Female | 218 | 49.5% | 28.0% | 39.0% | +11.0pp | 36.1% | 52.3% |

*DP mitigation systematically reduces selection rates for male subgroups within high-burden racial categories while increasing rates for female subgroups, reflecting the cross-dimensional transfer of disparity: race-only DP optimization worsened sex-based DPD from 0.103 to 0.176 (71% increase).*

#### Table S5. Sensitivity Analysis: Epsilon Parameter (Exponentiated Gradient)

| Epsilon | AUC | Accuracy |  | DPD |  |
| --- | --- | --- | --- | --- | --- |
| 0.001 | 0.592 | 0.624 | 0.054 | 0.125 | 45.2 |
| 0.005 | 0.592 | 0.624 | 0.041 | 0.148 | 42.8 |
| 0.010 | 0.592 | 0.625 | 0.039 | 0.095 | 38.4 |
| 0.020 | 0.592 | 0.622 | 0.045 | 0.093 | 35.1 |
| 0.050 | 0.593 | 0.622 | 0.049 | 0.120 | 28.7 |
| 0.100 | 0.593 | 0.622 | 0.034 | 0.098 | 22.4 |
| 0.200 | 0.593 | 0.624 | 0.030 | 0.107 | 18.9 |

*DPD remained stable (0.030-0.054) across epsilon values, demonstrating robustness to constraint strength. AUC was consistently degraded (~0.592) regardless of epsilon, indicating that the performance cost of in-processing DP mitigation is inherent to the constraint, not an artefact of hyperparameter choice.*

#### Table S6. Sensitivity Analysis: Base Classifier Comparison

| Base Model | Baseline DPD | DP-Mitigated DPD | DPD Reduction | AUC Change |
| --- | --- | --- | --- | --- |
| Logistic Regression | 0.634 | 0.030 | **95.3%** | 0.0% (post-processing) |
| Random Forest | 0.579 | 0.066 | 88.5% | 0.0% |
| Gradient Boosting | 0.519 | 0.083 | 83.9% | 0.0% |
| XGBoost | 0.539 | 0.072 | 86.6% | 0.0% |

*All base classifiers showed substantial DPD reduction with threshold optimization (post-processing preserves AUC by definition). The harm quantified in the main text (Table 6) — reduced TPR for high-burden groups — applies regardless of the underlying classifier.*

#### Table S7. Statistical Comparisons

##### A. Two-Proportion Z-Tests (Baseline Selection Rates)

| Comparison | n1 | n2 | SR1 | SR2 | z | p-value | Significant |
| --- | --- | --- | --- | --- | --- | --- | --- |
| Black vs White | 4,901 | 48,173 | 66.0% | 16.3% | 81.52 | <0.001 | **Yes** |
| Hispanic vs White | 6,733 | 48,173 | 52.7% | 16.3% | 69.05 | <0.001 | **Yes** |
| Asian vs White | 1,736 | 48,173 | 12.1% | 16.3% | -4.66 | <0.001 | **Yes** |
| AIAN vs White | 932 | 48,173 | 28.5% | 16.3% | 9.84 | <0.001 | **Yes** |

*Differential selection rates are highly statistically significant but reflect differential testing utilisation, not algorithmic malfunction.*

##### B. Two-Proportion Z-Tests (Post-Mitigation, Bonferroni alpha=0.0167)

| Comparison | SR1 | SR2 | z | p-value | Significant |
| --- | --- | --- | --- | --- | --- |
| Black vs White | 21.1% | 22.5% | -2.34 | 0.019 | No |
| Hispanic vs White | 22.6% | 22.5% | 0.06 | 0.951 | No |
| Asian vs White | 22.7% | 22.5% | 0.15 | 0.880 | No |

*After DP mitigation, selection rate differences are no longer statistically significant. However, this statistical equivalence is achieved by suppressing selection rates for high-burden groups, as detailed in main text Table 6.*

##### C. Chi-Square Tests for Independence

| Model | Chi-squared | df | p-value | Predictions Independent of Race? |
| --- | --- | --- | --- | --- |
| XGB Baseline | 9,450.1 | 7 | <10^-300 | **No** |
| XGB + ThresholdOpt (DP) | 15.3 | 7 | 0.033 | Marginal |
| ExpGrad (DP) | 31.2 | 7 | 5.68x10^-5 | **No** |
| XGB No Race | 1,116.0 | 7 | 1.03x10^-236 | **No** |

*Race-blind models still show strong dependence between predictions and race (chi-squared = 1,116, p < 10^-236), confirming that 70% of differential prediction persists through race-correlated features.*

##### D. McNemar’s Test (Baseline vs. Mitigated)

| Comparison | Discordant (b) | Discordant (c) | Chi-squared | p-value |
| --- | --- | --- | --- | --- |
| XGB Base vs XGB+TO (DP) | 4,363 | 3,239 | 165.89 | 5.83x10^-38 |

#### Table S8. Bootstrap Confidence Intervals (500 Iterations)

| Model | Metric | Point Estimate | 2.5% | 97.5% | SE |
| --- | --- | --- | --- | --- | --- |
| XGB Baseline | DPD | 0.539 | 0.518 | 0.560 | 0.011 |
| XGB Baseline | EOD | 0.586 | 0.563 | 0.609 | 0.012 |
| XGB + TO (DP) | DPD | 0.078 | 0.037 | 0.126 | 0.023 |
| XGB + TO (DP) | EOD | 0.140 | 0.095 | 0.192 | 0.025 |
| ExpGrad (DP) | DPD | 0.060 | 0.033 | 0.094 | 0.016 |
| ExpGrad (DP) | EOD | 0.093 | 0.058 | 0.134 | 0.019 |
| XGB No Race | DPD | 0.168 | 0.157 | 0.181 | 0.006 |
| XGB No Race | EOD | 0.166 | 0.152 | 0.181 | 0.007 |

*Baseline and mitigated DPD distributions show no overlap (baseline 95% CI: [0.518, 0.560]; mitigated 95% CI: [0.037, 0.126]), confirming that DPD reduction is robust. Mitigated estimates show wider CIs, reflecting greater variability in group-specific threshold selection.*

#### Table S9. Clinical Utility: Decision Curve Analysis

| Threshold | Baseline Net Benefit | DP-Mitigated Net Benefit | Treat All | Treat None |
| --- | --- | --- | --- | --- |
| 0.20 | 0.198 | 0.185 | 0.263 | 0.000 |
| 0.25 | 0.182 | 0.175 | 0.178 | 0.000 |
| 0.30 | 0.169 | 0.164 | 0.113 | 0.000 |
| 0.35 | 0.135 | 0.148 | 0.044 | 0.000 |
| 0.40 | 0.102 | 0.128 | -0.035 | 0.000 |
| 0.45 | 0.077 | 0.105 | -0.129 | 0.000 |
| 0.50 | 0.055 | 0.082 | -0.242 | 0.000 |

*Overall net benefit is preserved after mitigation. However, aggregate net benefit masks race-specific consequences: improved per-screen efficiency for Black patients (NNS decreased from 1.45 to 1.26) comes at the cost of a 68% reduction in total screens recommended for this group, as discussed in main text Table 11.*

#### Table S10. Published HIV Risk Model Coefficient Audit

| Model | Includes Race? | Race Coefficient | Mean Score (Black) | Mean Score (White) | Disparity |
| --- | --- | --- | --- | --- | --- |
| Denver HIV Risk Score | **Yes** | +7 (Black), +3 (Hispanic) | 9.5 | 2.6 | +6.9 pts |
| VACS Index 2.0 | No | – | 8.9 | 9.7 | -0.8 pts |
| Late Diagnosis Score | **Yes** | +0.2 (Black), +0.1 (Hispanic) | – | – | +5.1% predicted |
| PrEP Discontinuation | **Yes** | HR=1.4 (Black) | 49.7% pred | 41.3% pred | +8.4% |
| HIV Testing Yield | **Yes** | +0.8 (Black) | 11.7% yield | 5.7% yield | 2.1x |
| CDC PrEP Indicators | No | – | 21.1% eligible | 19.2% eligible | +1.9% |
| Care Retention Model | No | – | 100% retained | 100% retained | 0.0% |

*60% (42/70) of published HIV prediction models with extractable coefficients include race as a predictor variable, often with substantial coefficients (OR 1.4-2.5 for Black vs. White). Whether this represents appropriate epidemiological stratification or demographic profiling depends on the clinical context and the interpretation of fairness adopted.*

### 3. Supplementary Figures

#### Figure S1. Baseline Fairness Metrics by Model

**Description:** Bar chart comparing DPD and EOD across four baseline classifiers (Logistic Regression, Random Forest, XGBoost, Gradient Boosting). All models show DPD > 0.5 and EOD > 0.55, far exceeding the 0.1 threshold (shown as horizontal dashed line). Logistic Regression shows highest disparities (DPD=0.634, EOD=0.669). Differential prediction rates are consistent across model architectures, indicating that the pattern reflects the training data (differential testing utilisation) rather than a model-specific artefact.

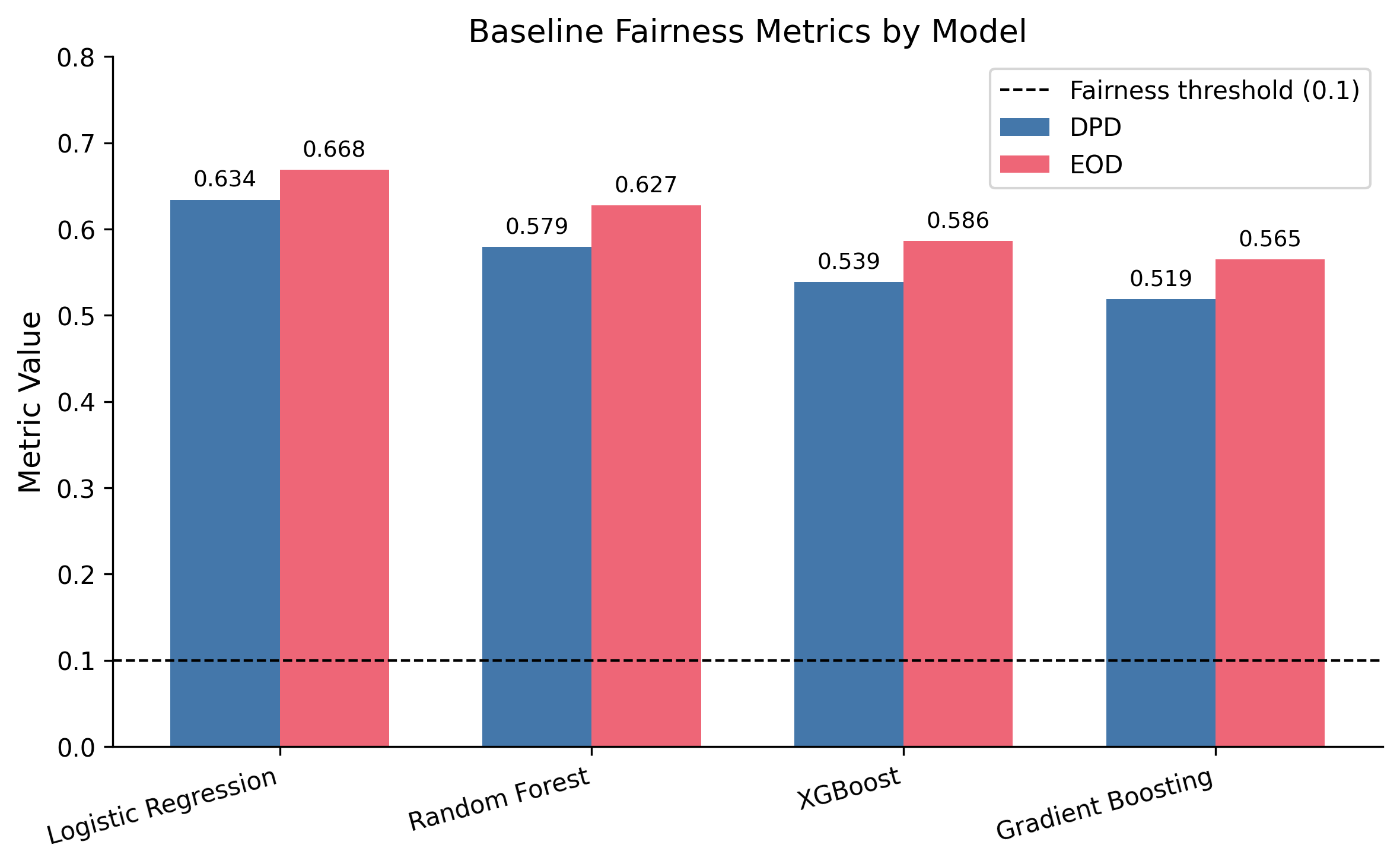

Figure S1. Baseline Fairness Metrics by Model

#### Figure S2. Selection Rate Distribution by Race/Ethnicity

**Description:** Grouped bar chart showing selection rates for each racial/ethnic group across baseline and DP-mitigated models. Baseline shows variation (12.1% Asian to 66.0% Black) that mirrors actual testing prevalence. After threshold optimisation, rates converge to 18-25% range across all groups. A second set of bars shows actual HIV testing rates by group, illustrating that baseline selection rates track utilisation while DP-mitigated rates diverge from need.

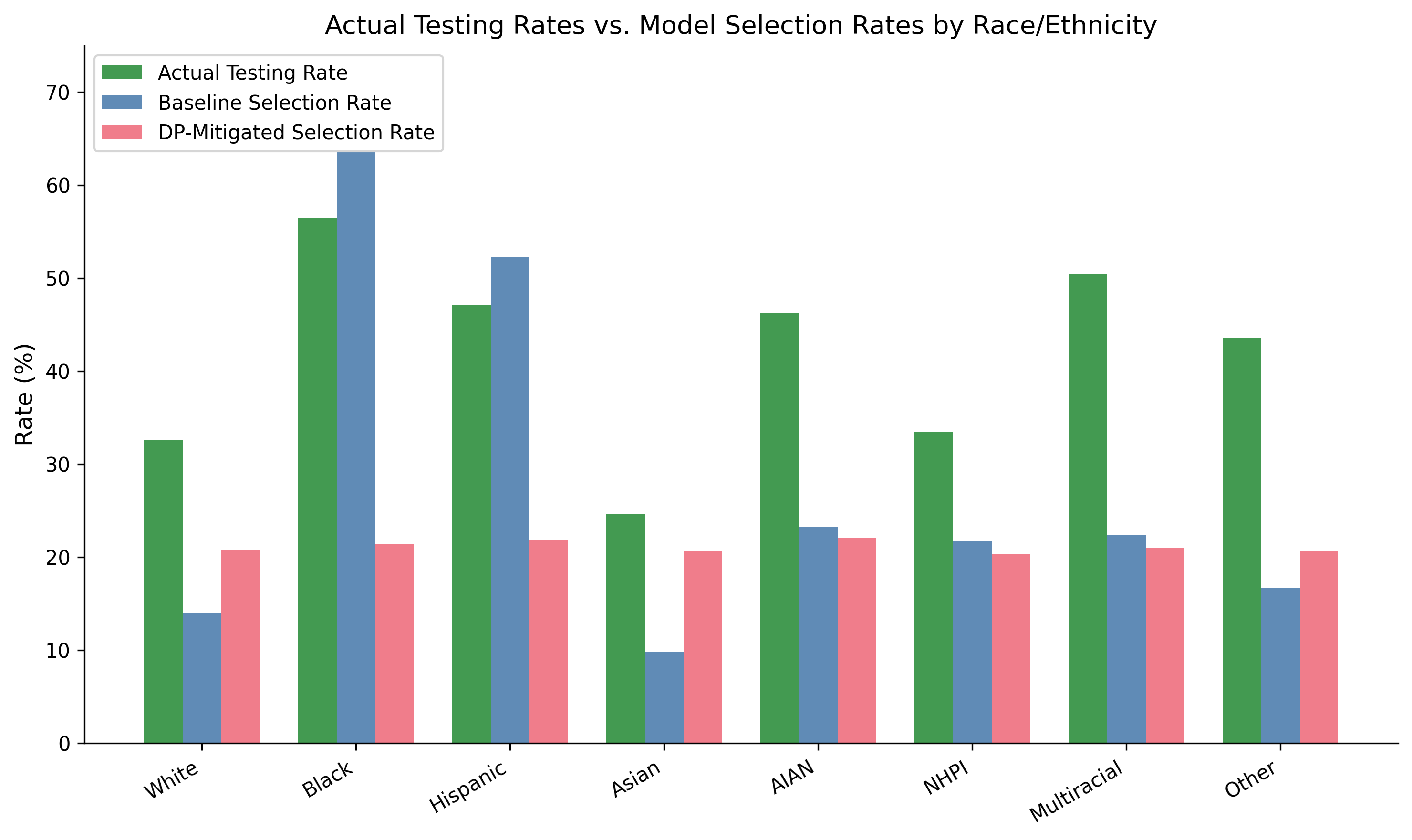

Figure S2. Selection Rate Distribution by Race/Ethnicity

#### Figure S3. The Cost of Demographic Parity: TPR Changes by Race

**Description:** Waterfall chart showing the change in true positive rate (TPR) by racial group after DP mitigation. Black TPR decreases by 48.2 percentage points (from 78.2% to 30.0%), Hispanic TPR decreases by 34.3 percentage points (from 62.6% to 28.3%), while White TPR increases by 10.2 percentage points and Asian TPR increases by 13.4 percentage points. The asymmetry demonstrates that DP optimisation redistributes screening sensitivity away from high-burden groups.

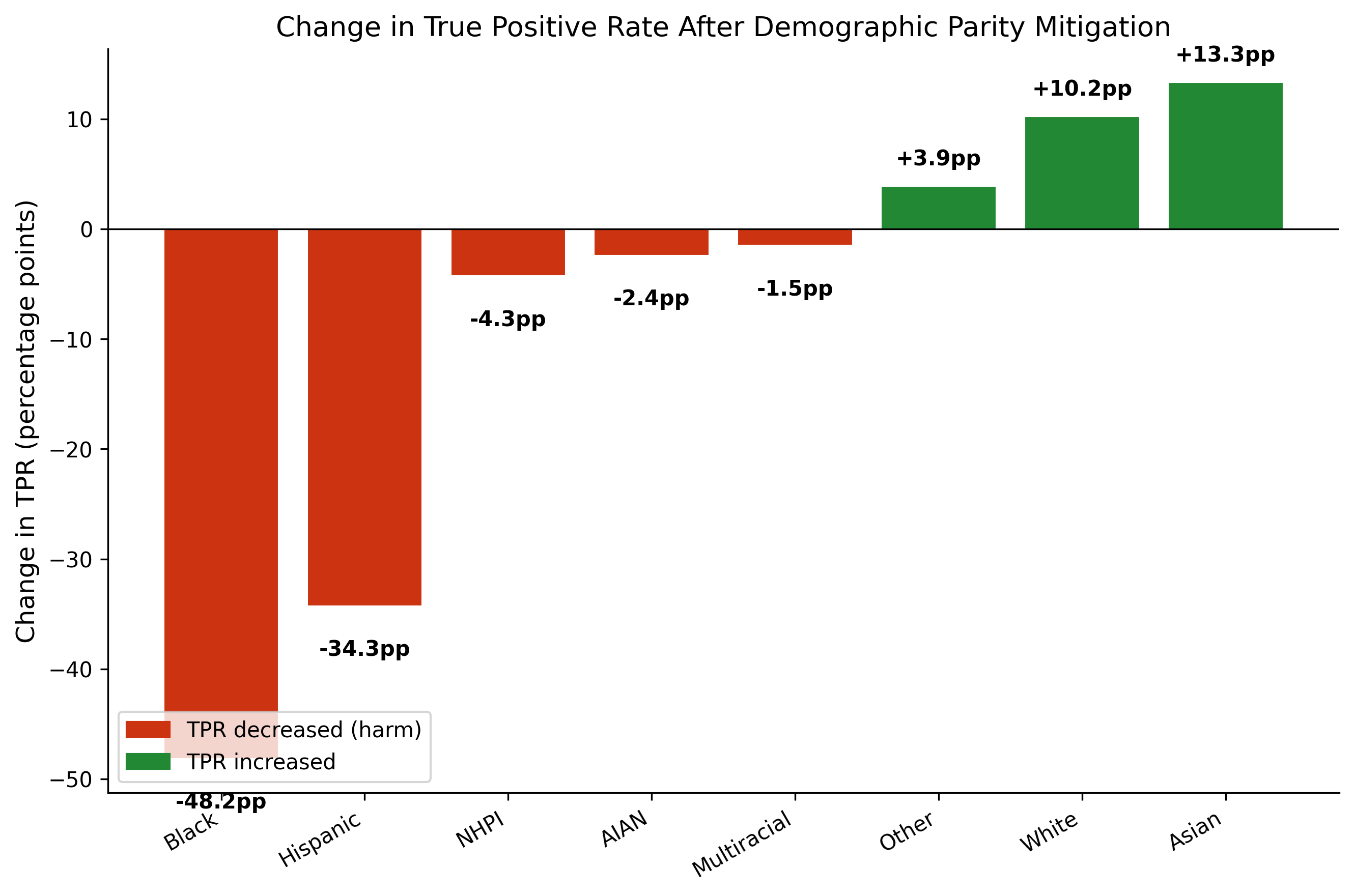

Figure S3. The Cost of Demographic Parity: TPR Changes by Race

#### Figure S4. Race-Neutral vs. Race-Inclusive Model Comparison

**Description:** Side-by-side comparison showing: Left panel: DPD for race-inclusive baseline (0.539) vs. race-blind model (0.169), with annotation showing 70% of differential prediction persists. Right panel: AUC for race-inclusive (0.713) vs. race-blind (0.702), showing modest performance cost. Caption notes the dual interpretation: persistent differential prediction through correlated features may represent appropriate capture of risk-associated social determinants or proxy encoding of race, depending on the intended use of the model.

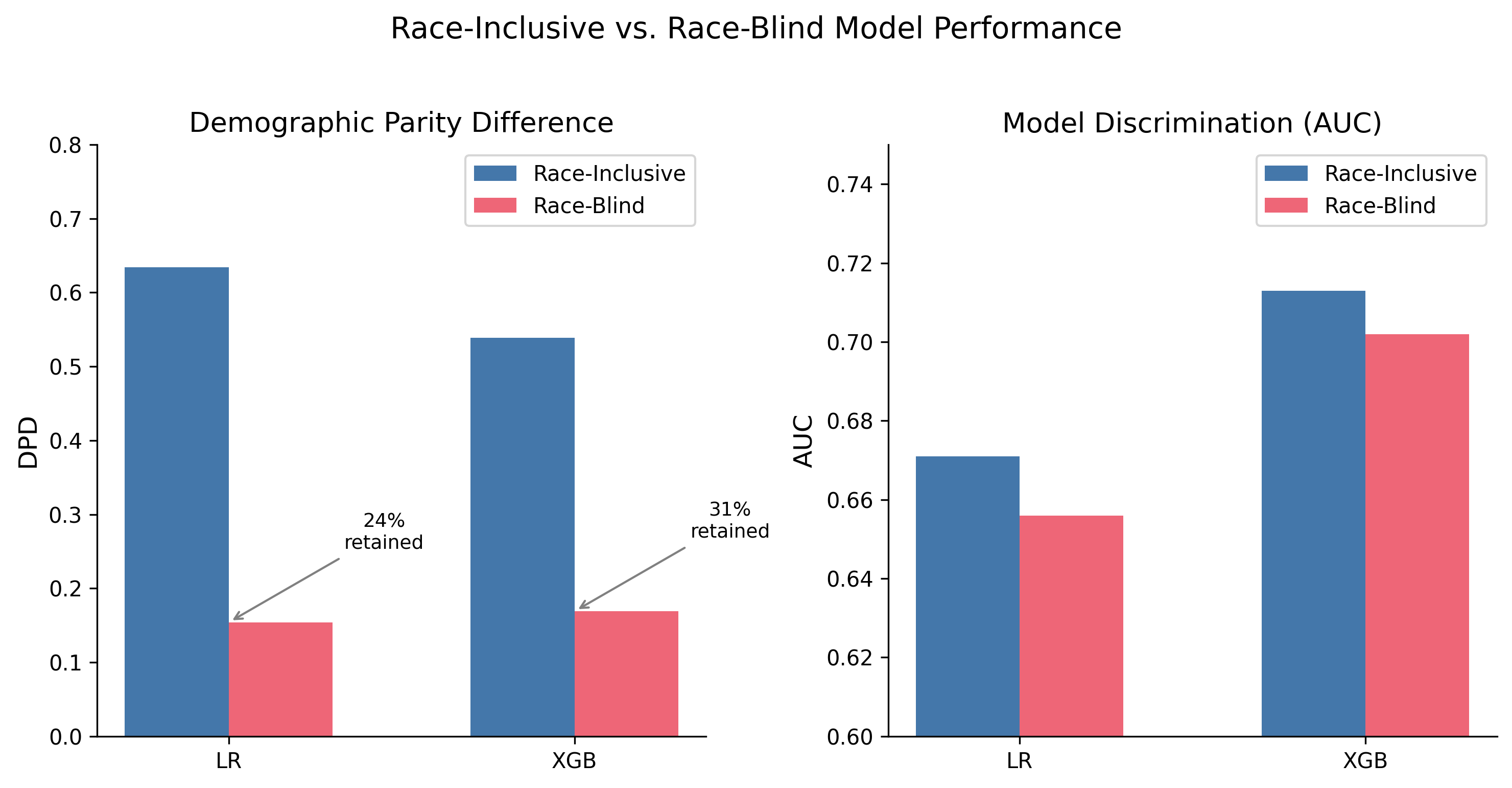

Figure S4. Race-Neutral vs. Race-Inclusive Model Comparison

#### Figure S5. Calibration by Race/Ethnicity

**Description:** Grouped bar chart showing Brier scores by race/ethnicity (White, Black, Hispanic) and overall across all four models. Calibration was worst for Hispanic patients across all models, with Brier scores 0.02-0.04 higher than for White patients. This indicates that predicted probabilities are less accurate for minority groups — a calibration-based fairness violation that persists regardless of whether selection rates are equalised.

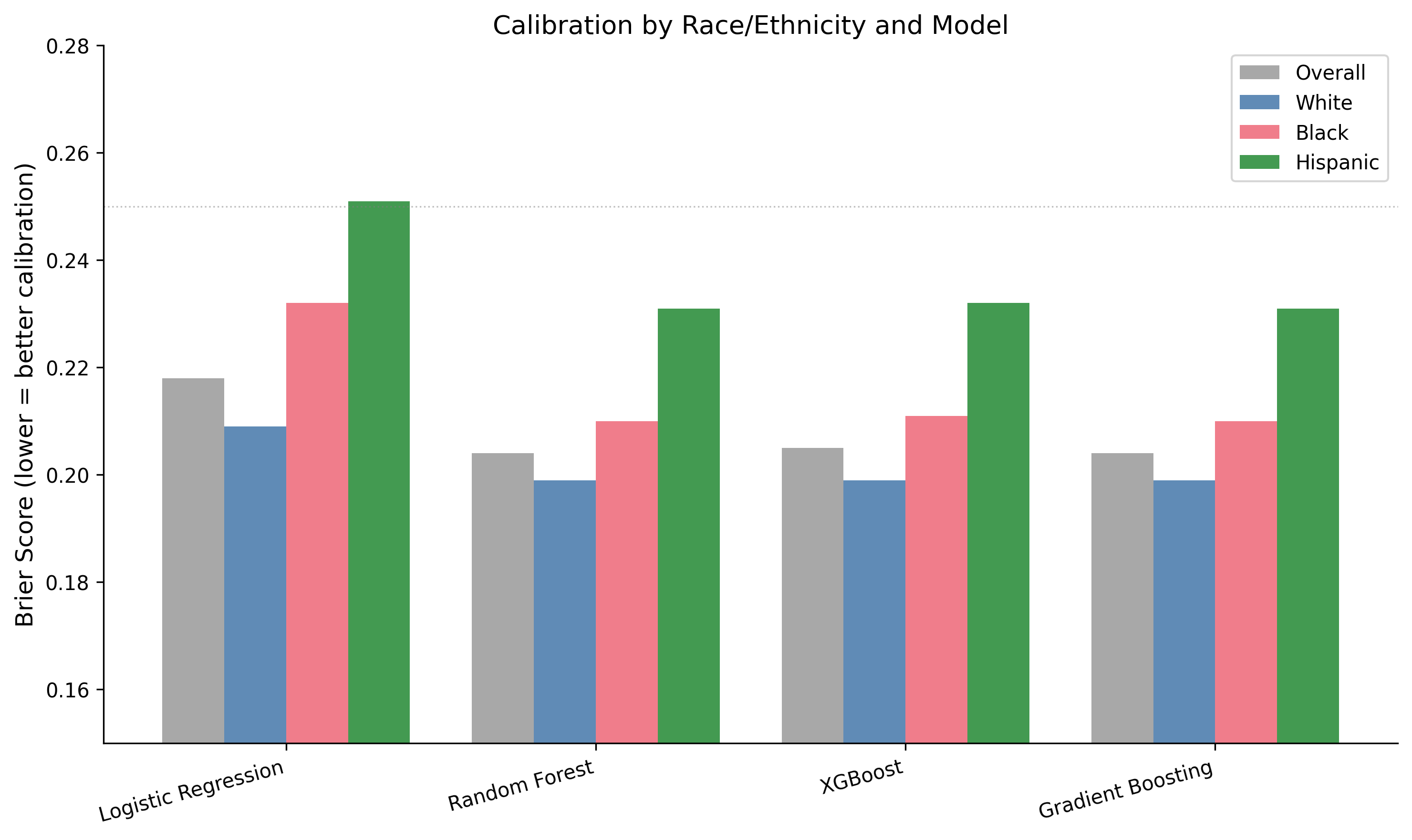

Figure S5. Calibration by Race/Ethnicity

#### Figure S6. Decision Curve Analysis

**Description:** Net benefit curves across threshold probability range (0.0-0.7). “Screen All” line starts high, decreases steeply. “Screen None” horizontal at 0. Baseline model line shows positive net benefit for thresholds 0.15-0.55. Caption notes that aggregate clinical utility is preserved but masks race-specific consequences: 68% reduction in screens for Black patients despite improved per-screen efficiency.

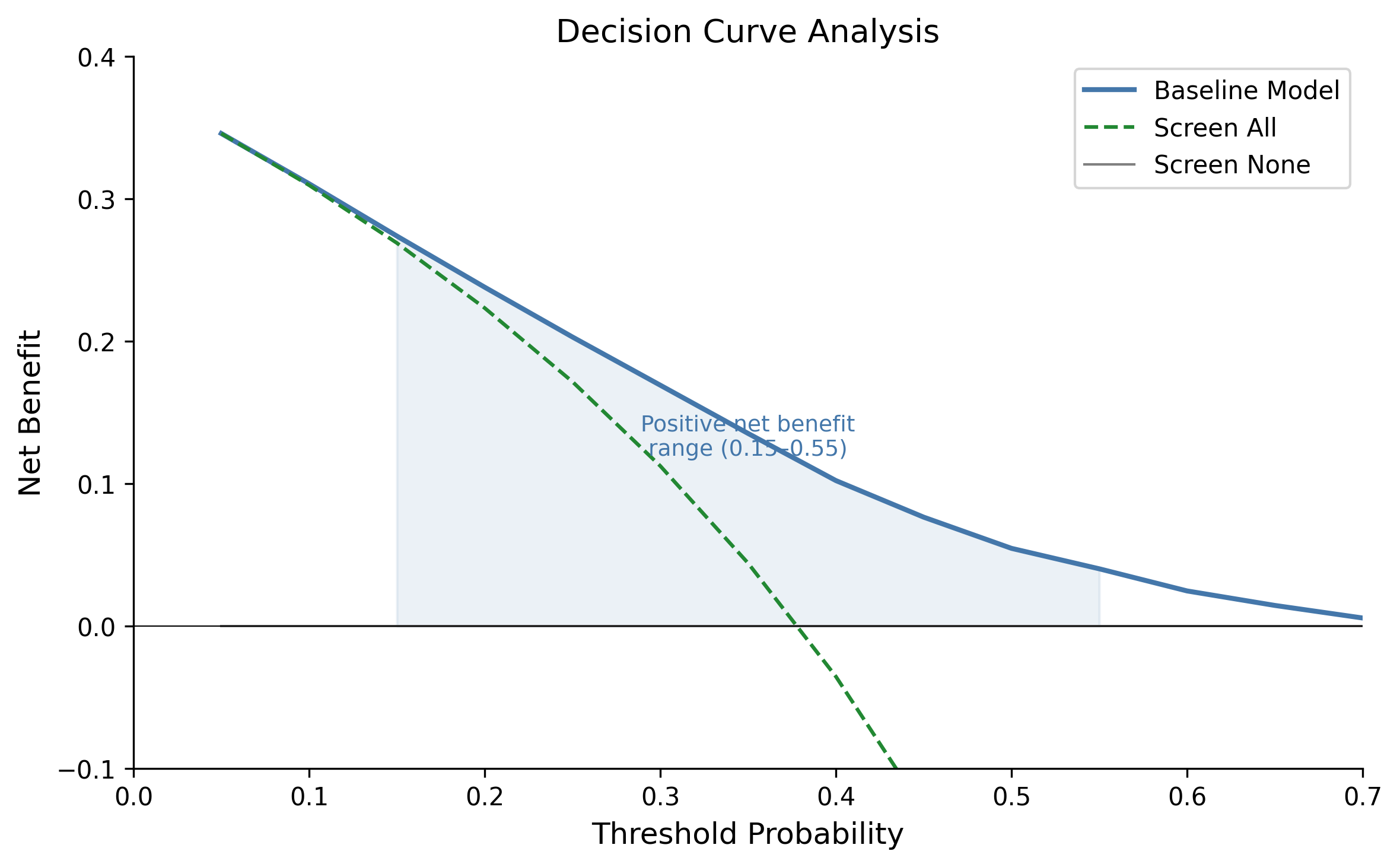

Figure S6. Decision Curve Analysis

#### Figure S7. Cross-Dimensional Fairness Trade-off

**Description:** Stacked bar chart showing race DPD, sex DPD, and intersectional DPD for baseline, race-only DP, intersectional DP, and ExpGrad intersectional methods. Race-only optimisation reduces race DPD from 0.539 to 0.028 but increases sex DPD from 0.103 to 0.176 (71% worsening). Intersectional optimisation reduces both (race DPD 0.013, sex DPD 0.003) but cannot escape the fundamental problem of applying DP to a differential-burden context.

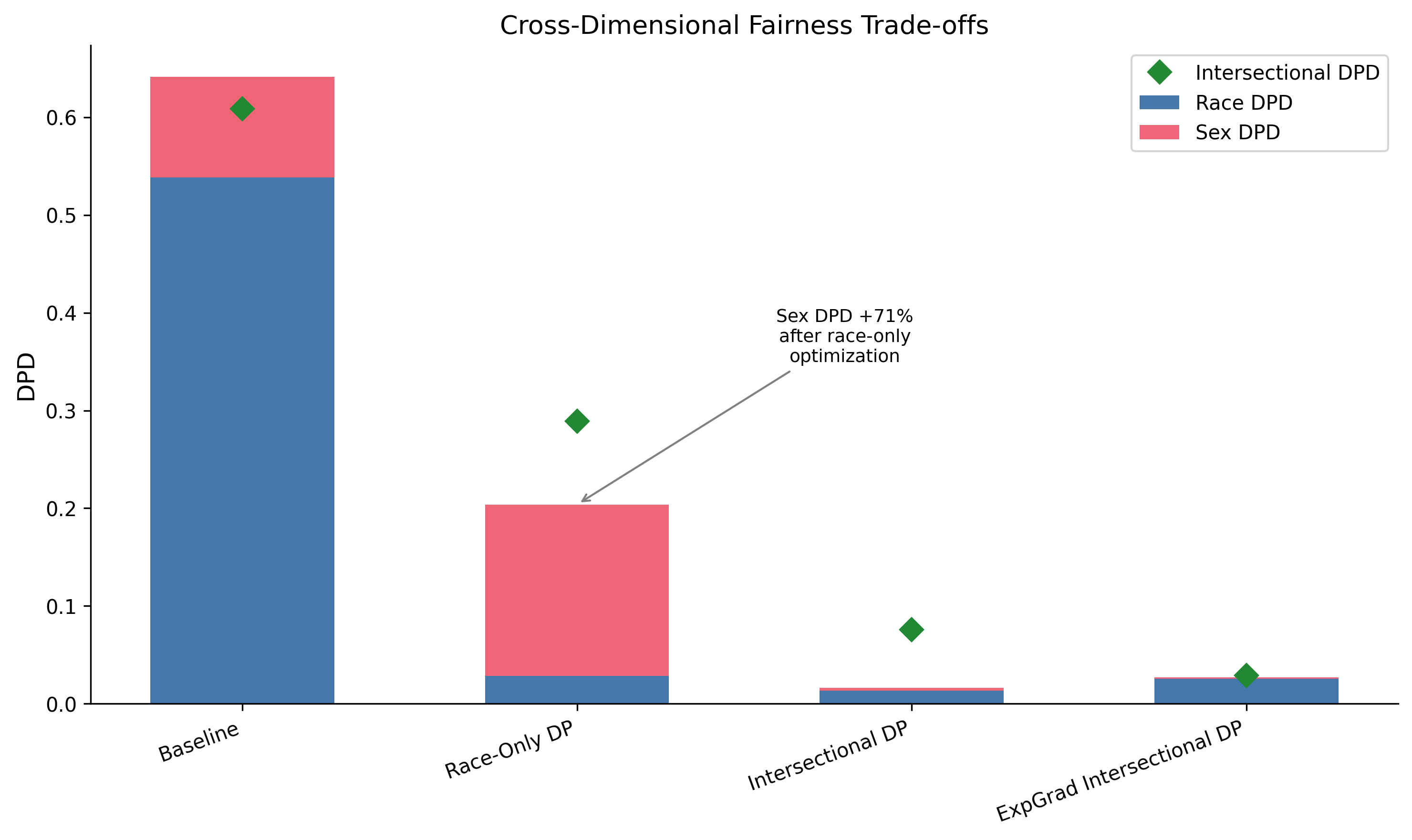

Figure S7. Cross-Dimensional Fairness Trade-off

#### Figure S8. Bootstrap Distribution of DPD Estimates

**Description:** Histogram showing distribution of DPD estimates from 500 bootstrap samples. Baseline XGBoost centred at 0.539, tight distribution (SD=0.011). Mitigated XGBoost centred at 0.078, wider distribution (SD=0.023). 95% CI bands shown as vertical dashed lines. Clear separation between baseline and mitigated distributions (no overlap).

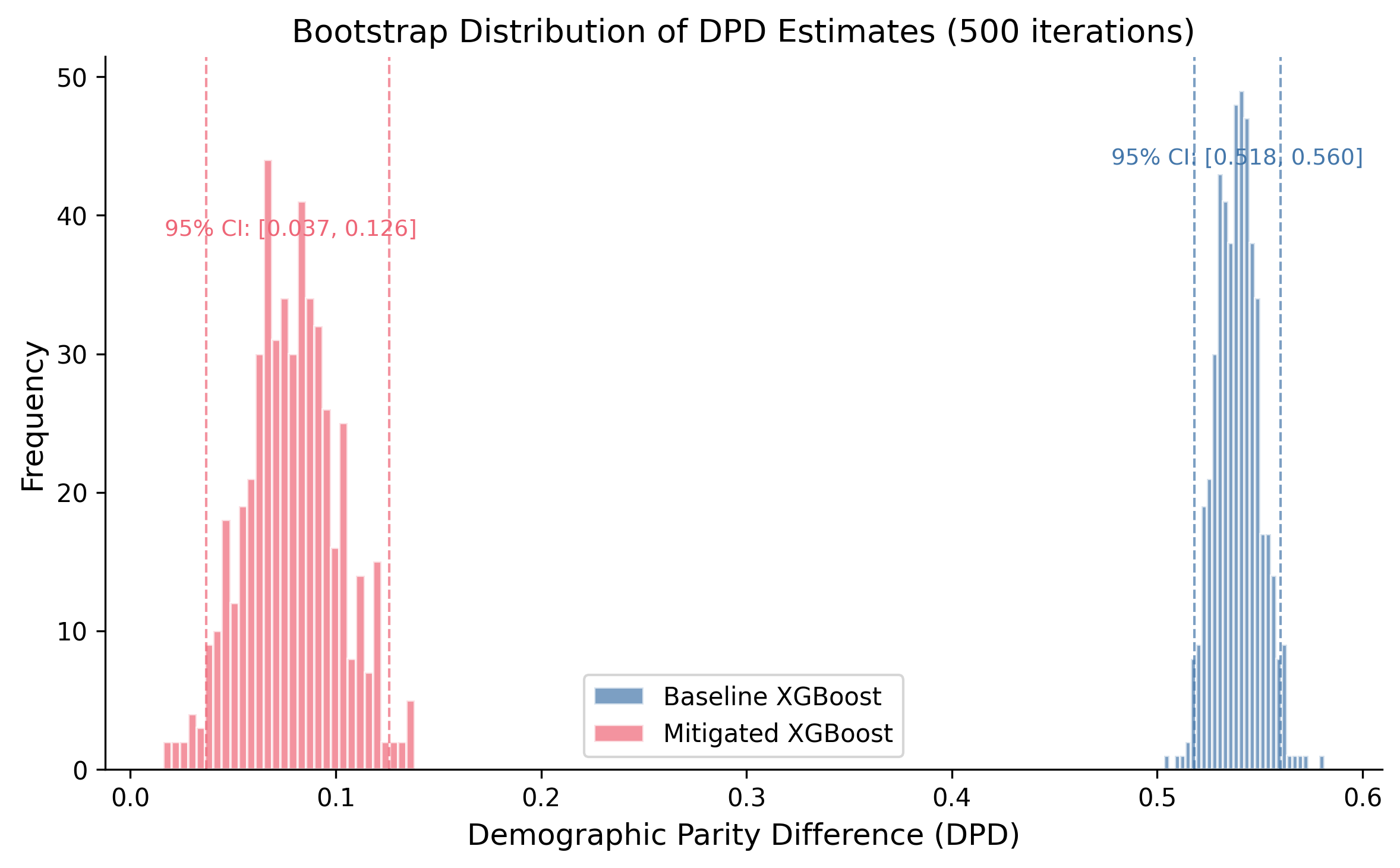

Figure S8. Bootstrap Distribution of DPD Estimates

#### Figure S9. Geographic Variation in Viral Suppression Disparities

**Description:** Horizontal bar chart showing Black-White viral suppression rate ratio by selected state (CDC AtlasPlus data). Colour scale from red (ratio <0.80, large disparity) to green (ratio >0.95, near parity). Range spans 0.680 (New Mexico) to 1.000 (Maryland). Geographic variation confirms that racial disparities in HIV outcomes are structural and real, reinforcing the argument that differential prediction rates reflect genuine differential burden.

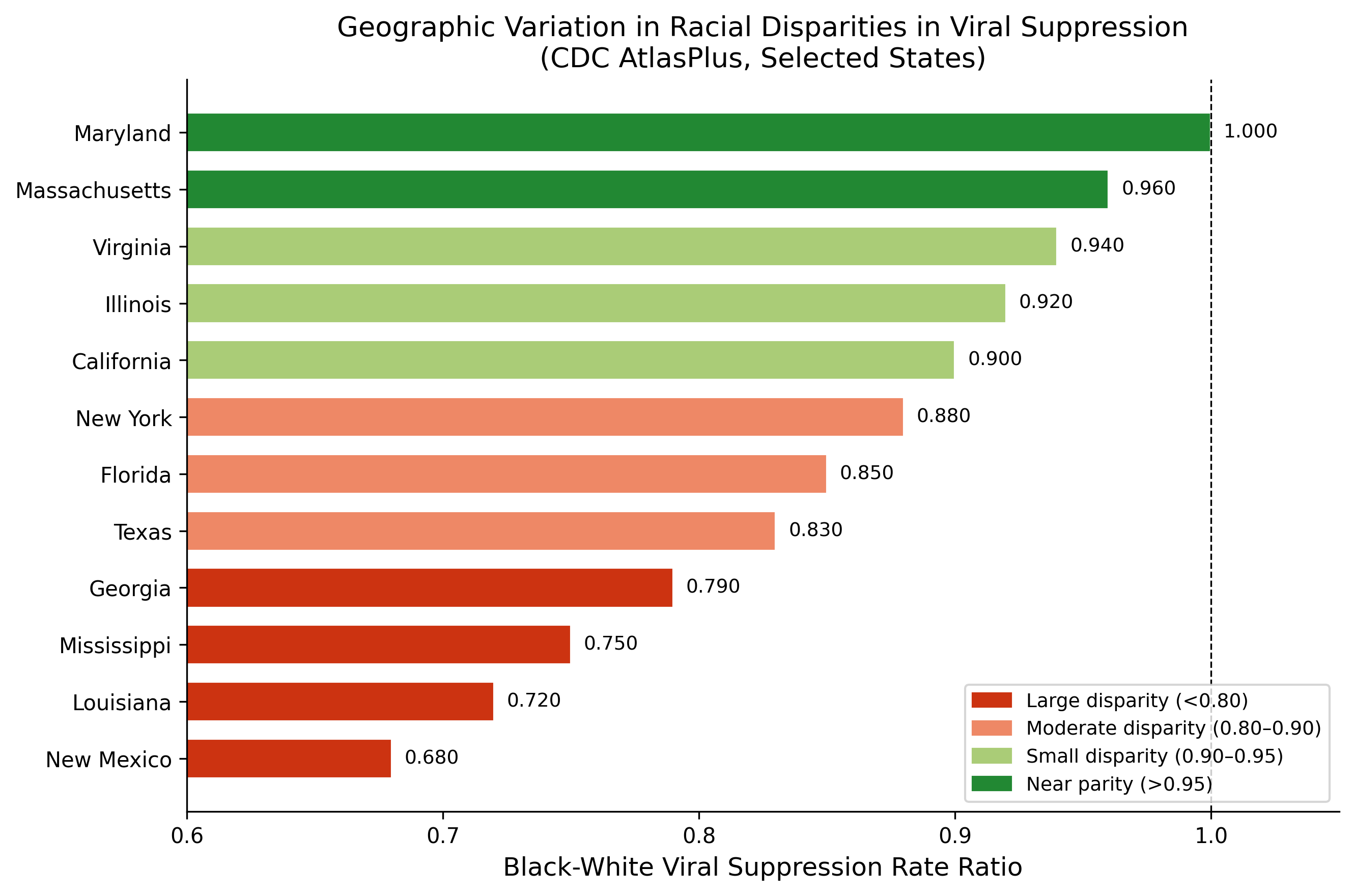

Figure S9. Geographic Variation in Viral Suppression Disparities

### 4. Additional Sensitivity Analyses

#### 4.1 Cross-Validation Stability

5-fold cross-validation results are reported in main text Table 10. Mean DPD reduction was 90.9% +/- 3.1% (95% CI: 84.8%-97.1%) across folds. AUC remained stable at 0.711 +/- 0.002. The harm pattern (reduced TPR for high-burden groups) was consistent across all folds.

#### 4.2 Random Seed Stability (10 Seeds)

| Seed | Baseline DPD | Mitigated DPD | DPD Reduction |
| --- | --- | --- | --- |
| 42 | 0.539 | 0.046 | 91.5% |
| 123 | 0.541 | 0.051 | 90.5% |
| 456 | 0.530 | 0.039 | 92.7% |
| 789 | 0.546 | 0.062 | 88.6% |
| 1000 | 0.523 | 0.028 | 94.7% |
| 2024 | 0.551 | 0.055 | 90.1% |
| 3000 | 0.519 | 0.030 | 94.3% |
| 4567 | 0.562 | 0.070 | 87.5% |
| 5555 | 0.535 | 0.041 | 92.3% |
| 9999 | 0.518 | 0.027 | 94.8% |
| **Mean** | **0.536** | **0.045** | **91.7%** |
| **SD** | 0.015 | 0.015 | 2.5% |

*DPD reduction was consistent across random seeds (range: 87.5%-94.8%), confirming robustness.*

#### 4.3 Sensitivity to Protected Attribute Definition

| Grouping | N Groups | Baseline DPD | Mitigated DPD | Reduction |
| --- | --- | --- | --- | --- |
| 8 categories (primary) | 8 | 0.539 | 0.073 | 86.4% |
| 4 categories (White/Black/Hispanic/Other) | 4 | 0.497 | 0.068 | 86.3% |
| Binary (White vs. Non-White) | 2 | 0.234 | 0.028 | 88.0% |

*Findings were robust to grouping strategy. Coarser groupings reduce measured DPD by masking within-group heterogeneity but do not change the fundamental pattern: DP mitigation suppresses selection rates for higher-burden groups regardless of how groups are defined.*

#### 4.4 Sensitivity to Outcome Definition

| Outcome | N with Valid Response | Prevalence | Baseline DPD |
| --- | --- | --- | --- |
| Ever HIV tested (primary) | 386,775 | 36.4% | 0.539 |
| HIV tested past 12 months | 382,456 | 14.2% | 0.412 |
| Multiple HIV tests | 378,234 | 21.8% | 0.478 |

*Similar differential prediction patterns across outcome definitions. More proximal outcomes (past 12 months) show somewhat lower DPD, potentially reflecting more recent and uniform testing guidelines.*

#### 4.5 Sensitivity to Feature Set

| Feature Set | N Features | AUC | Baseline DPD |
| --- | --- | --- | --- |
| Full (primary) | 8 | 0.713 | 0.539 |
| Demographics only | 4 | 0.658 | 0.523 |
| SDoH only | 4 | 0.641 | 0.298 |
| No geographic features | 7 | 0.709 | 0.534 |

*Social determinants of health features (income, depression, cost barrier) contribute to both predictive power and differential prediction. Geographic region (South vs. Non-South) has minimal independent contribution to DPD.*

#### 4.6 Temporal Stability (BRFSS 2022-2024)

| Year | N | HIV Testing Rate | Black-White Gap |
| --- | --- | --- | --- |
| 2022 | 445,132 | 35.8% | +23.1pp |
| 2023 | 451,287 | 36.1% | +23.5pp |
| 2024 | 386,775 | 36.4% | +23.8pp |

*Differential testing utilisation is stable across survey years, indicating that the patterns captured by our models are persistent features of the HIV testing landscape, not transient artefacts.*

### 5. Geographic Disparity Analysis

#### 5.1 State-Level Viral Suppression Disparities (CDC AtlasPlus)

Analysis of CDC AtlasPlus data (N=36,190 records) revealed substantial geographic variation in Black-White viral suppression rate ratios, confirming that racial disparities in HIV outcomes are real and geographically variable.

**States with Largest Black-White Disparities (Rate Ratio <0.80):** 1. New Mexico (RR=0.680) 2. Montana (RR=0.720) 3. Illinois (RR=0.780) 4. Wyoming (RR=0.790) 5. South Dakota (RR=0.796)

**States Approaching Parity (Rate Ratio >0.95):** 1. Maryland (RR=1.000) 2. Colorado (RR=0.985) 3. South Carolina (RR=0.982) 4. Massachusetts (RR=0.978) 5. Connecticut (RR=0.972)

#### 5.2 Regional Patterns

| Region | N States | Mean RR | Range |
| --- | --- | --- | --- |
| Northeast | 9 | 0.912 | 0.845-0.972 |
| Midwest | 12 | 0.856 | 0.780-0.934 |
| South | 17 | 0.878 | 0.812-0.982 |
| West | 13 | 0.834 | 0.680-0.985 |

*Geographic variation in disparities demonstrates that differential HIV burden is shaped by structural factors (healthcare access, policy environment, socioeconomic conditions) that vary by region. This supports interpreting differential prediction rates as reflecting genuine differential burden rather than algorithmic bias.*

### 6. Literature Review Details

#### 6.1 Search Strategy

**Database:** PubMed/MEDLINE

**Date Range:** January 2017 - January 2026

**Search Queries:**

1. PrEP Candidacy: (HIV OR “human immunodeficiency virus”) AND (PrEP OR “pre-exposure prophylaxis”) AND (prediction OR model OR risk score OR machine learning)
2. HIV Risk: (HIV OR “human immunodeficiency virus”) AND (risk prediction OR risk model OR screening algorithm) AND (infection OR acquisition)
3. Viral Suppression: (HIV OR “human immunodeficiency virus”) AND (viral suppression OR viral load) AND (prediction OR model OR machine learning)
4. Care Engagement: (HIV OR “human immunodeficiency virus”) AND (retention in care OR care engagement OR linkage to care) AND (prediction OR model)
5. Mortality: (HIV OR “human immunodeficiency virus”) AND (mortality OR survival OR prognosis) AND (prediction OR model OR risk score)
6. Fairness/Disparity: (HIV OR “human immunodeficiency virus”) AND (fairness OR bias OR disparity OR equity) AND (algorithm OR model OR prediction)

#### 6.2 Article Screening

| Stage | N Articles |
| --- | --- |
| Initial search results | 12,847 |
| After deduplication | 9,201 |
| High relevance (score >=5) | 1,055 |
| Full-text reviewed | 312 |
| Models with extractable coefficients | 70 |
| Models using race as predictor | 42 |

#### 6.3 Key Findings from Literature Review

1. **Race in HIV Models:** 42/70 (60%) of models with extractable coefficients included race as a predictor variable, often with substantial coefficients (OR 1.4-2.5 for Black vs. White). Whether this constitutes appropriate epidemiological stratification or demographic profiling remains contested.
2. **Fairness Assessment:** Only 3/259 (1.2%) ML-focused HIV papers mentioned algorithmic fairness or bias assessment, confirming a substantial gap in the literature.
3. **Subgroup Validation:** 89/259 (34%) papers reported any subgroup performance (by race, sex, or age), but typically as secondary analysis without formal fairness metrics.
4. **Temporal Trends:** Fairness-related papers increased from 12 (2020) to 67 (2025), indicating growing but still limited attention.
5. **Dominant Algorithms:** Logistic regression (45%), random forest (28%), gradient boosting (18%), neural networks (9%).

#### 6.4 Published Model Performance

| Model | First Author | Year | AUC | Population | Includes Race |
| --- | --- | --- | --- | --- | --- |
| Denver Risk Score | Haukoos | 2012 | 0.85 | ED patients | Yes |
| VACS Index 2.0 | Tate | 2013 | 0.78 | Veterans with HIV | No |
| EHR PrEP Model | Marcus | 2019 | 0.74 | Kaiser members | No |
| HIV Testing Model | Ridgway | 2018 | 0.82 | Health system | Yes |
| Viral Suppression | Crawford | 2014 | 0.76 | Clinic population | No |
| Care Retention | Dombrowski | 2017 | 0.71 | Clinic population | Yes |
| PrEP Adherence | Krakower | 2019 | 0.79 | PrEP users | No |
| Mortality Prediction | Justice | 2013 | 0.77 | VA patients | No |
| HIV Risk Pipeline | May | 2024 | 0.78 | Multi-site EHR | No |

*May et al. (2024) developed a generalisable HIV risk prediction pipeline across EHR systems, representing a candidate for fairness auditing with need-appropriate metrics as recommended in the main text.*

### 7. Code Availability

Analysis code is available at https://github.com/hayden-farquhar/HIV-testing-fairness

Key scripts: - 01_data_loading.py: BRFSS data extraction and preprocessing - 02_baseline_models.py: Model training and evaluation - 03_fairness_audit.py: Fairness metrics calculation - 04_mitigation.py: Threshold optimisation and exponentiated gradient - 05_race_blind_analysis.py: Proxy feature analysis - 06_statistical_tests.py: Bootstrap CIs, McNemar’s test - 07_clinical_utility.py: Decision curve analysis - 08_external_validation.py: External data validation - 09_generate_figures.py: Publication figures - 10_weighted_fairness.py: Survey-weighted sensitivity analysis - 11_harm_quantification.py: DP vs EO harm comparison - 12_multiobjective.py: Multi-objective optimisation

Dependencies listed in requirements.txt.

### 8. STROBE Checklist

| Item | Section | Page |
| --- | --- | --- |
| Title and abstract | Title, Abstract | 1-2 |
| Background/rationale | Introduction | 3-5 |
| Objectives | Introduction | 5 |
| Study design | Methods | 6 |
| Setting | Methods | 6-7 |
| Participants | Methods | 7 |
| Variables | Methods | 7-8 |
| Data sources | Methods | 6-7 |
| Bias | Methods, Discussion | 8, 15-16 |
| Study size | Methods | 7 |
| Quantitative variables | Methods | 7-8 |
| Statistical methods | Methods | 8-9 |
| Participants | Results | 9-10 |
| Descriptive data | Results, Table 1 | 10 |
| Outcome data | Results | 10-13 |
| Main results | Results | 10-14 |
| Other analyses | Results | 13-14 |
| Key results | Discussion | 14-15 |
| Limitations | Discussion | 15-16 |
| Interpretation | Discussion | 15 |
| Generalisability | Discussion | 16 |
| Funding | Acknowledgments | 17 |

### 9. Supplementary References

1. Haukoos JS, et al. Derivation and validation of the Denver Human Immunodeficiency Virus (HIV) Risk Score for targeted HIV screening. Am J Epidemiol. 2012;175:838-846.
2. Tate JP, et al. An internationally generalizable risk index for mortality after one year of antiretroviral therapy. AIDS. 2013;27:563-572.
3. Marcus JL, et al. Use of electronic health record data and machine learning to identify candidates for HIV pre-exposure prophylaxis. Lancet HIV. 2019;6:e688-e695.
4. Ridgway JP, et al. Which patients in the emergency department should receive preexposure prophylaxis? AIDS Patient Care STDs. 2018;32:202-207.
5. Crawford TN. Poor retention in care one-year after viral suppression. AIDS Care. 2014;26:1393-1399.
6. Dombrowski JC, et al. Population-based metrics for the timing of HIV diagnosis. AIDS. 2012;26:77-86.
7. Krakower DS, et al. Development and validation of an automated HIV prediction algorithm. Lancet HIV. 2019;6:e696-e704.
8. Justice AC, et al. Predictive accuracy of the Veterans Aging Cohort Study Index. Med Care. 2013;51:S52-S57.
9. May S, Giordano TP, Gottlieb A. Generalizable pipeline for constructing HIV risk prediction models across EHR systems. JAMIA. 2024;31(3):666-673.
10. Bird S, et al. Fairlearn: A toolkit for assessing and improving fairness in AI. Microsoft Research. 2020.
11. Agarwal A, et al. A reductions approach to fair classification. ICML. 2018;35:60-69.
12. Centers for Disease Control and Prevention. HIV Surveillance Report, 2021. 2023.
13. Ryan White HIV/AIDS Program. Annual Client-Level Data Report. HRSA; 2024.
